# Development and Validation of the Hypertension Population Risk Tool: A Population-Based Diagnostic Algorithm for Canadians

**DOI:** 10.64898/2025.12.10.25342022

**Authors:** Rafidul Islam, Tracey Bushnik, Manish M. Sood, Monica Taljaard, Finlay A. McAlister, Juan Li, Douglas G. Manuel

## Abstract

**Objectives:** Effective, equitable hypertension prevention requires an understanding of which populations are at risk. We aimed to develop and validate the Hypertension Population Risk Tool (HTNPoRT) - a diagnostic model derived with only readily available data, suitable for individual screening and population health planning.

**Methods:** We analyzed data from the Canadian Health Measures Survey (cycles 1–6, 2007–2019). The study included community-dwelling respondents aged 20–79 years. The primary outcome was hypertension, defined as measured systolic/diastolic blood pressure of 140/90 mm Hg or current antihypertensive medication use. Sex-specific logistic regression models were developed using 16 predictors, including 4 sociodemographic, 3 psychosocial, 2 health status, 5 health behavioural, and 2 chronic condition variables. The model was fully prespecified, including the stepdown procedure to derive parsimonious models.

**Results:** Of 19,643 participants, 5,152 (26.2%) had hypertension. The final models included age, body mass index, diabetes, and family history of hypertension. Optimism-corrected c-statistics were 0.86 (95% CI: 0.85–0.87) for men and 0.88 (95% CI: 0.87–0.88) for women. Calibration showed relative differences between observed and predicted risk of 1.02% (men) and 1.41% (women), and consistent performance across 179 of 181 policy-relevant subgroups. Predicted hypertension risk in Canada varied but rose markedly with older age, diabetes, and obesity.

**Conclusions:** HTNPoRT is a well-performing predictive algorithm that relies only on minimal non-invasive, self-reported data. It is suitable for both individual risk screening and population-level surveillance to inform hypertension prevention strategies targeting both the general population and high-risk groups.

## Introduction

Hypertension affects about 1 in 4 Canadians and is the world’s leading risk factor for preventable cardiovascular disease and all-cause mortality (Leung et al., 2019; Murray et al., 2020). However, reducing this burden is challenged by the asymptomatic nature of the condition; many individuals remain undiagnosed or unaware of their status (Inoue, 2025). Diagnostic risk algorithms can bridge this gap by first identifying high-risk individuals who may have undiagnosed hypertension, thereby reinforcing the need for confirmed clinical blood pressure measurement. Beyond this immediate screening function, accurate risk classification can guide prevention strategies by facilitating individual decision-making and supporting the identification of high-risk subpopulations to inform the evaluation and design of preventive interventions tailored to population needs (Manuel et al., 2012).

Despite these potential benefits, there is a lack of accessible tools that can provide this baseline risk assessment. To be widely usable for screening or surveillance, a risk assessment approach must rely on non-invasive, self-reportable, and population-representative predictors. However, most existing prediction algorithms are not aligned with these requirements. They often use predictors inaccessible for individual self-assessment and cumbersome for population-level planning, such as genetic markers, biomarkers, and clinically obtained measures (Fava et al., 2013; Kanegae et al., 2018; Niiranen et al., 2016). Furthermore, they are typically developed using ethnically homogenous cohorts, limiting their generalizability to diverse populations needed for effective health planning (Fava et al., 2013; Kanegae et al., 2018).

Our objective was to develop and validate a diagnostic algorithm - the Hypertension Population Risk Tool (HTNPoRT) - that the public and health planners can use to classify the risk of hypertension. We specifically set out to develop the algorithm using multiethnic, population-based survey data and readily available, self-reported measures to facilitate screening reinforcement and inform prevention strategies.

## Methods

### Study design

We developed and validated sex-specific HTNPoRT algorithms using a registered protocol (OSF.io, 10.17605/OSF.IO/V27MW) (Islam et al., 2024). Our reporting followed the TRIPOD-AI guidelines (Collins et al., 2024). The statistical approach to developing the model followed recommendations by both Steyerberg and Harrell (Harrell, 2016; Steyerberg, 2009). The Ottawa Health Science Network Research Ethics Board granted approval for the research (20240693-01H). There were minor deviations from the protocol to improve the model’s final performance and usability; a detailed list and justification for each deviation is provided in Appendix 1.

### Data source

Data for the study included the first six available cycles of the Canadian Health Measures Survey (CHMS), collected from March 2007 to December 2019. The CHMS is a recurring, cross-sectional survey conducted by Statistics Canada designed to produce national-level health data. It uses a stratified, multi-stage sampling design to select a representative sample of community-dwelling individuals aged 3 to 79 living in the 10 provinces (regardless of their medical access or blood pressure awareness). The survey excludes full-time members of the Canadian Forces and people living in the three territories, on reserves, in institutions, and in certain remote regions.

Data collection for the CHMS is a two-step process: an in-person household interview followed by a visit to a Mobile Examination Centre (MEC). The household interview gathers sociodemographic, health, and lifestyle information. At the MEC, a series of direct physical measurements are performed, including standardized blood pressure readings according to a published protocol. Information on prescription medication use, including antihypertensive drugs, is collected via inventory during both the household interview and the MEC visit (Statistics Canada, 2018). The combined response rate for respondents aged 20 to 79 in the six cycles was 50.6%. Further details on accessing the CHMS data are available in Appendix 2.

The study population consisted of community-dwelling Canadians aged 20 to 79 in the CHMS (N = 19,853). Respondents were excluded if they were pregnant or had missing values for the hypertension outcome, yielding a final sample size of 19,643 (9,633 males and 10,010 females).

### Outcome

The study outcome was hypertension status, determined by direct blood pressure measurements and a medication inventory. At the MEC, six blood pressure readings were taken from seated, unattended respondents using an automated BpTRU™ BPM-300 device. Following the CHMS protocol, the first reading was discarded, and the average of the final five readings was used.

These mean values were adjusted using a standard formula to approximate manual auscultatory readings (Bryan et al., 2010; Leung et al., 2019; Myers et al., 2008). The CHMS blood pressure measurement protocol is described more in Appendix 3. Respondents were classified as hypertensive if they met any of the following criteria:

1. A mean adjusted systolic blood pressure (SBP) of ≥ 140 mm Hg or a mean adjusted diastolic blood pressure (DBP) of ≥ 90 mm Hg. For respondents with diabetes or chronic kidney disease, a lower threshold of SBP ≥ 130 mm Hg or DBP ≥ 80 mm Hg was used.
2. Reported taking antihypertensive medication in the past month for the purpose of lowering blood pressure.

### Predictors

Seventeen candidate predictors were selected for the initial model based on review of established hypertension risk factors and consultation with knowledge users in clinical hypertension and public health (Table 1). These predictors were grouped into five domains: sociodemographic measures, psychosocial measures, health statuses, health behaviours, and chronic conditions. All predictor variables were cleaned, derived, and harmonized across the six CHMS cycles using functions from the chmsflow R package, a newly developed package to support open and reproducible harmonization across CHMS cycles (Islam et al., 2025). All variables with missing data were imputed once to create one imputed dataset, using a model derived from the multiple imputation using chained equations (MICE) method (Buuren & Groothuis-Oudshoorn, 2011). Detailed variable definitions and derivations are provided in Appendix 3 and the analytical code.

**Table 1.**
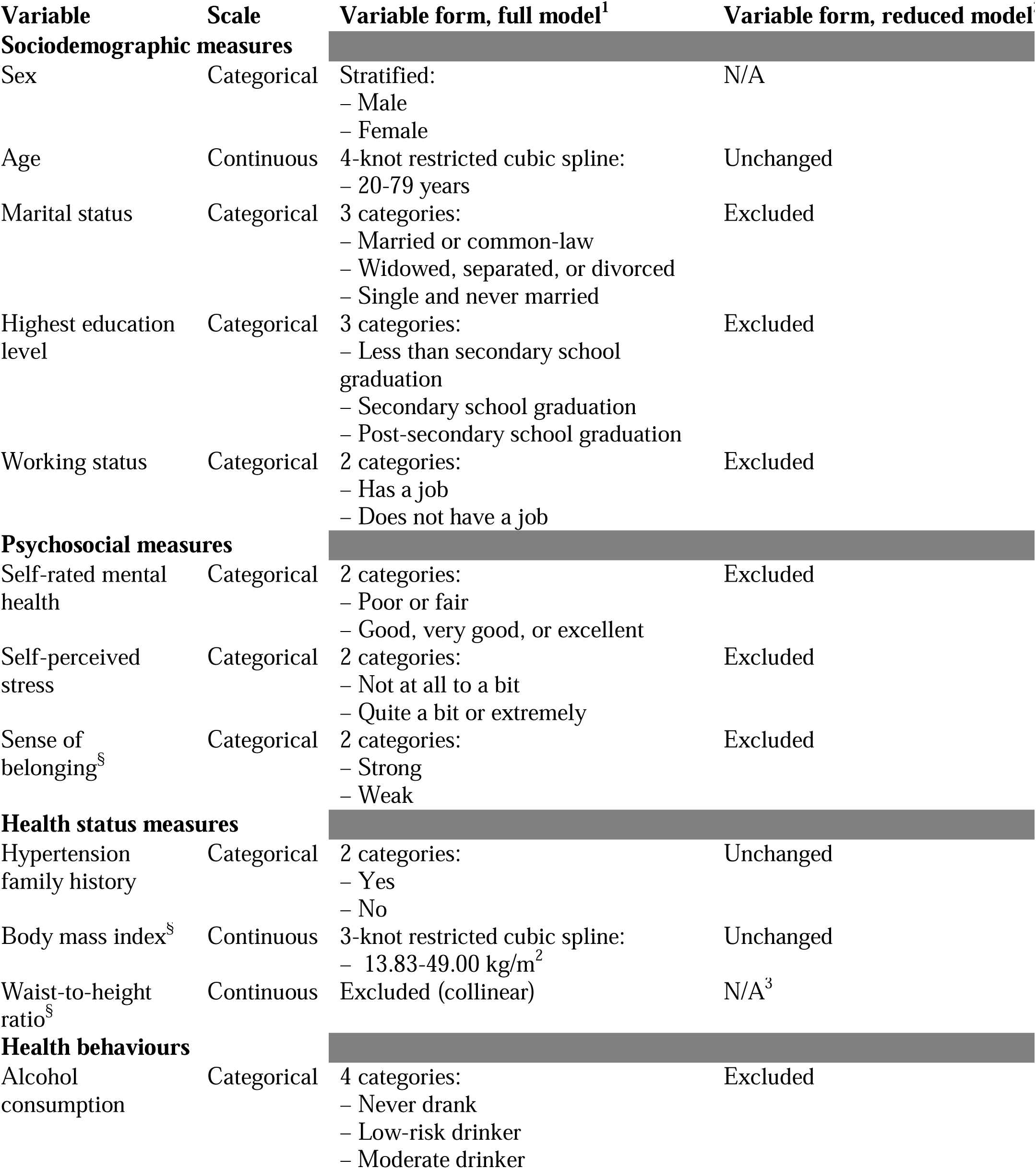
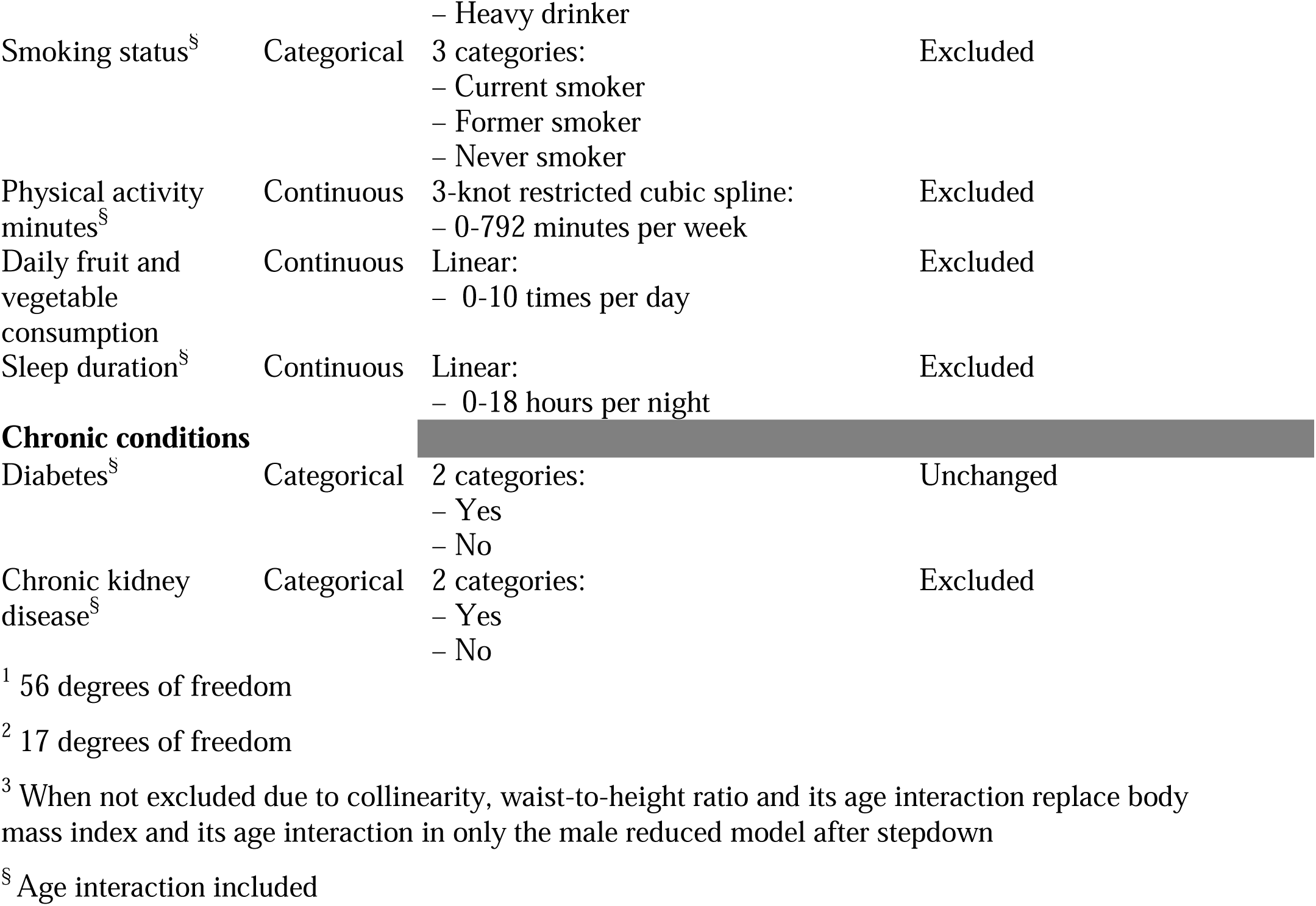
Predictor variables for the Hypertension Population Risk Tool (HTNPoRT)

Predictor data were drawn from three sources: the household questionnaire, direct physical measurements, and laboratory tests collected at the MEC. Direct physical measurements included height, weight, and waist circumference, which were all used to calculate body mass index (BMI) and waist-to-height ratio. Laboratory test measures included blood glucose and serum creatinine, which were used to derive the presence of diabetes and chronic kidney disease, respectively (Charles et al., 2024; Leung et al., 2019).

### Statistical analysis

Sex-specific logistic regression models were developed. All analyses incorporated CHMS survey weights for population-level inference. Continuous predictors were modelled with restricted cubic splines when there were known non-linear relationships; otherwise, they were modelled as linear terms. Knots for restricted cubic splines were placed at fixed quantiles of the distribution as described by Harrell (Harrell, 2016). The models incorporated two-way interactions between age and clinically relevant variables identified in the literature.

#### Model development and specification

An initial sex-specific “full model” was fitted using all pre-specified predictors and interactions (Table 1). To create a more practical model for end-users, a parsimonious “reduced model” was derived from each full model using a stepdown approach. This process sequentially removed predictors with the least contribution until the model’s R² was 95% of the full model’s R² (Ambler et al., 2002; Harrell, 2016).

#### Assessment of model performance

To compare the full and reduced models for each sex, we used an internal bootstrap approach. The models were refitted and evaluated on 1,000 bootstrap samples, each randomly drawn with replacement from the original sample from which they were derived. The models were also validated within their original derivation sample. Sensitivity analyses were performed to assess if model performance was affected by either dropping respondents with missing data, deriving models from four other imputed datasets generated with the same MICE imputation model, leaving skewness, re-including possible collinear predictors, using linear interactions, deriving hypertension with unadjusted blood pressures, excluding hypertensives classified on medications alone, or using 130/80 mm Hg blood pressure cutoffs to define hypertension for all respondents.

Assessment of overall model performance across the derivation and the bootstrap samples was detailed, focusing on measures of predictive accuracy, discrimination, and calibration. Predictive accuracy was gauged by Nagelkerke’s R² and the Brier score (Steyerberg et al., 2010).

Discrimination, or the model’s ability to distinguish between those who have the outcome and those who do not, was appraised via the concordance statistic (c-statistic) whose optimism was quantified using the 1,000 bootstrap samples of internal validation (Harrell, 2016; Steyerberg et al., 2010). Calibration, or the agreement between predicted and observed risk, was assessed by comparing observed and predicted hypertension probabilities, calibration slopes, and visual inspection of calibration plots fitted with Local Estimated Scatterplot Smoothing (Vergouwe et al., 2005). Calibration was also assessed across different subgroups important to clinicians and policymakers, including most of the model’s predictor variables (Table 1), as well as additional variables and alternate derivations of predictor variables. The clinically relevant standard of calibration was defined as a difference of less than 20% between observed and predicted estimates within subgroups with a hypertension prevalence of at least 5%. Centering variables around their means will facilitate recalibration in new populations.

#### Model presentation and implementation

All sex-specific logistic models were presented post-validation using beta coefficients. SHapley Additive exPlanation (SHAP) plots were used to show the ranked importance of risk factors in increasing or decreasing the predicted probability for a given individual, using one of the models. SHAP values provided an understanding of variable importance, accounting for non-linear effects and interactions (Ariza-Garzón et al., 2020). The current hypertension probability and the SHAP plot comprised the hypertension risk profile. Additionally, mean predicted probabilities were displayed for the overall population and select risk groups. To enhance accessibility to HTNPoRT models, we will render them executable via a user-friendly web application and an Excel-based calculator available here at https://osf.io/mjd7n/files/bgmcr.

## Results

### Participants

The final study population comprised 19,643 respondents, including 9,633 men and 10,010 women. Of this sample, 5,152 individuals (2,681 men and 2,471 women) were classified as having hypertension, corresponding to a weighted prevalence of 25% in men and 22% in women. This estimate can be interpreted as the prevalence of hypertension among Canadian adults from 2007 to 2019.

The median age was 46 years (IQR: 33-58) for men and 47 years (IQR: 34-59) for women. Detailed weighted characteristics of the study population are in Table 2. Most predictors had less than 2% missing data. Missing data was most prevalent for hypertension family history and sleep duration (30-38% missing), which were not collected in all CHMS cycles. A detailed breakdown of missingness for each predictor without weighting is shown in Appendix 4.

**Table 2.**
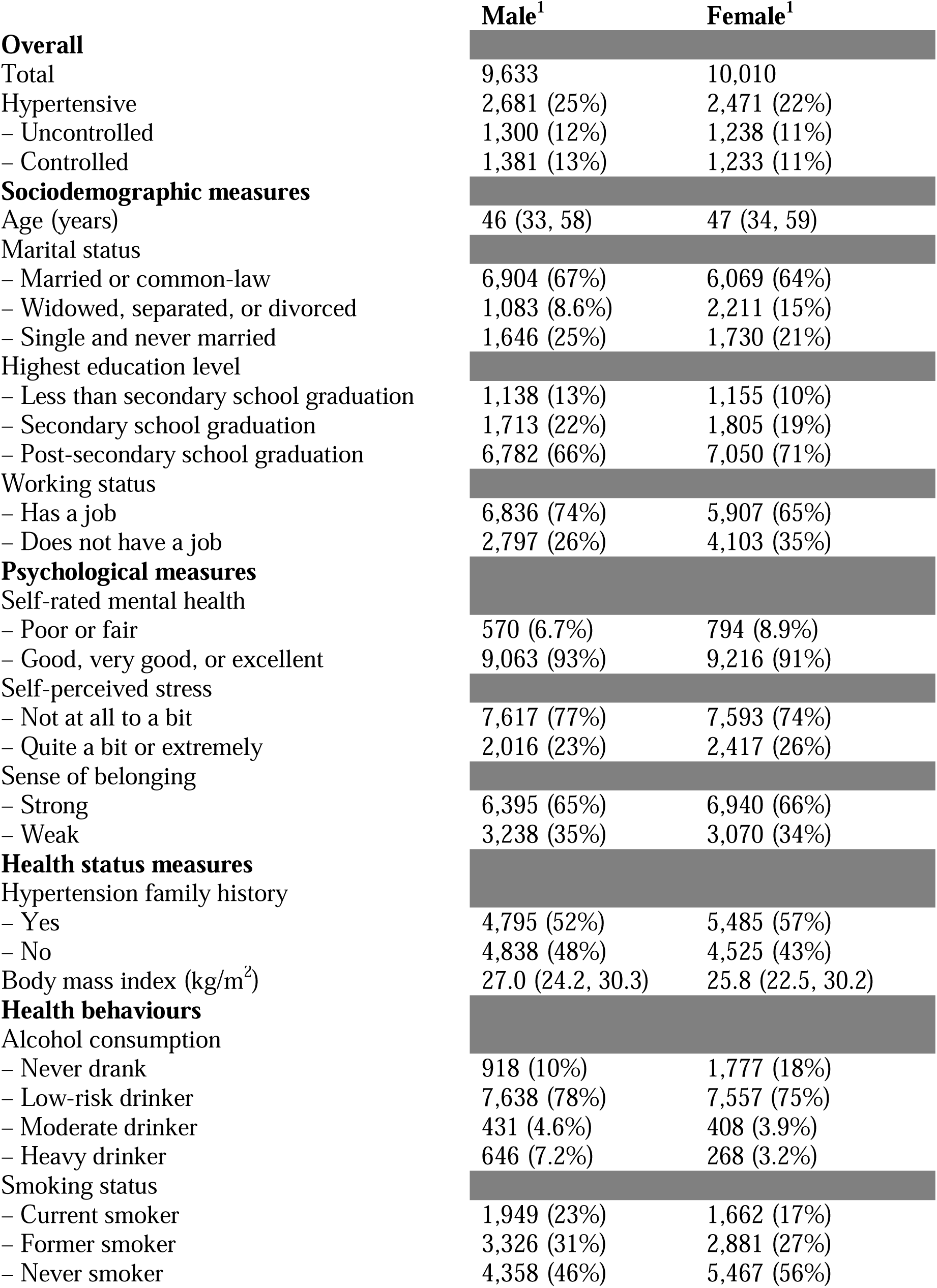
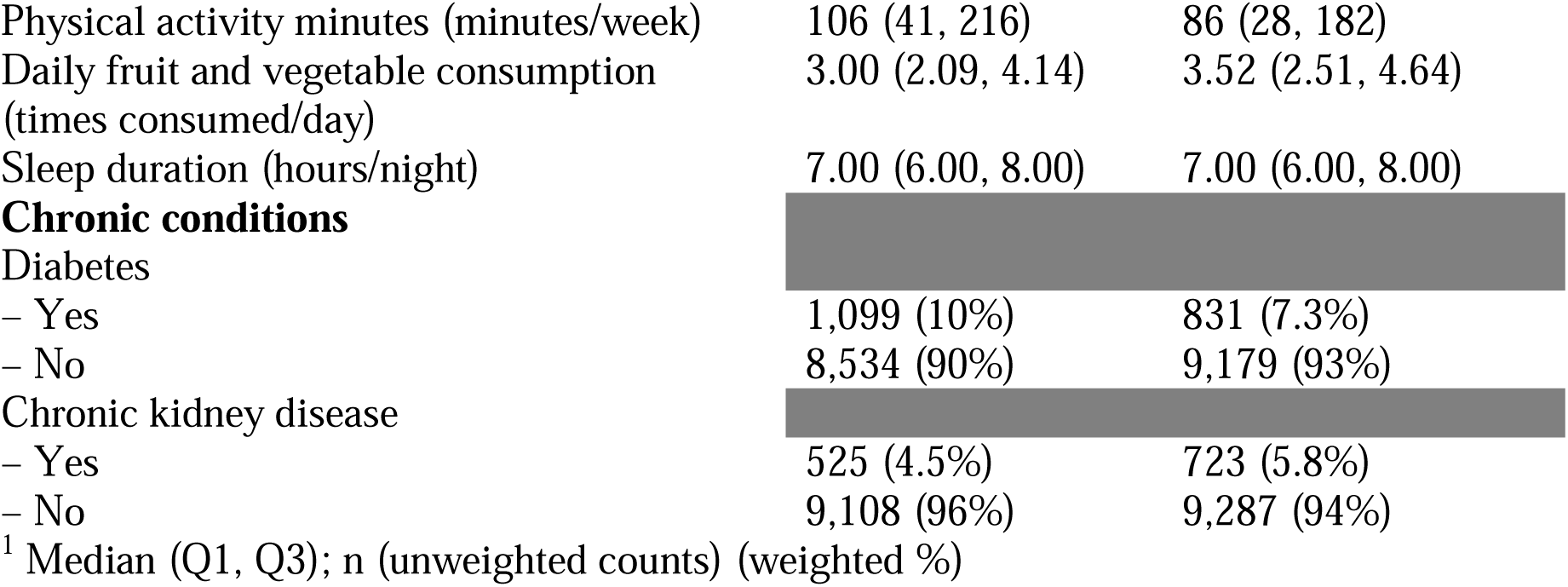
Weighted characteristics of the study populations for HTNPoRT.

### Model performance

The performance of two sets of sex-specific models was evaluated: a full model with 16 predictors and seven interactions (56 degrees of freedom), and a more parsimonious reduced model with four predictors and two interactions (17 degrees of freedom). Waist-to-height ratio was excluded because it was collinear with body mass index (variance inflation factor > 2.5). Model specifications and beta coefficients are available in Table 1 and Appendix 5, respectively.

#### Full models

Table 3 shows that the full models for both men and women were well able to discriminate those who have hypertension from those who do not (c-statistic across bootstrap samples, men: 0.87 (95% CI: 0.86-0.87); women: 0.88 (95% CI: 0.87-0.89)), with c-statistics remaining unchanged upon optimism correction. Therefore, predicted hypertension risk was concentrated in high-risk individuals (95:5 predicted probability ratios across bootstrap samples, men: 445.8, women: 714.5). Predicted probabilities of hypertension closely approximated observed proportions of hypertension within the overall population (observed v. predicted relative difference across bootstrap samples, men: 2.09%; women: 1.15%). Calibration slopes were near perfect (0.95 in men and 0.97 in women across bootstrap samples), with calibration plots for the full models being shown in Figure 1.

**Figure 1.**
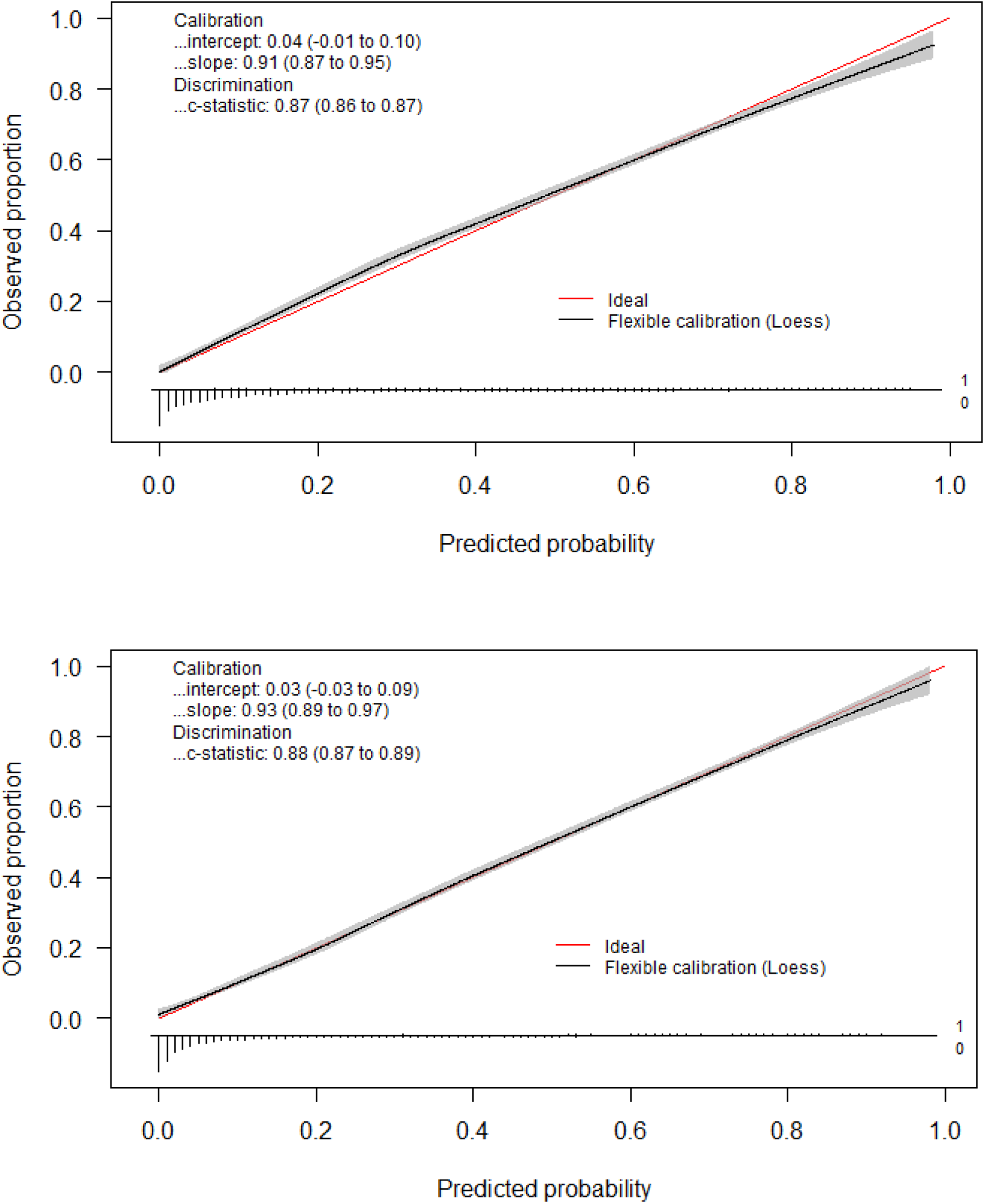
Calibration plots for full models; predicted hypertension probability versus observed hypertension proportion in men (top) and women (bottom)

**Table 3.**
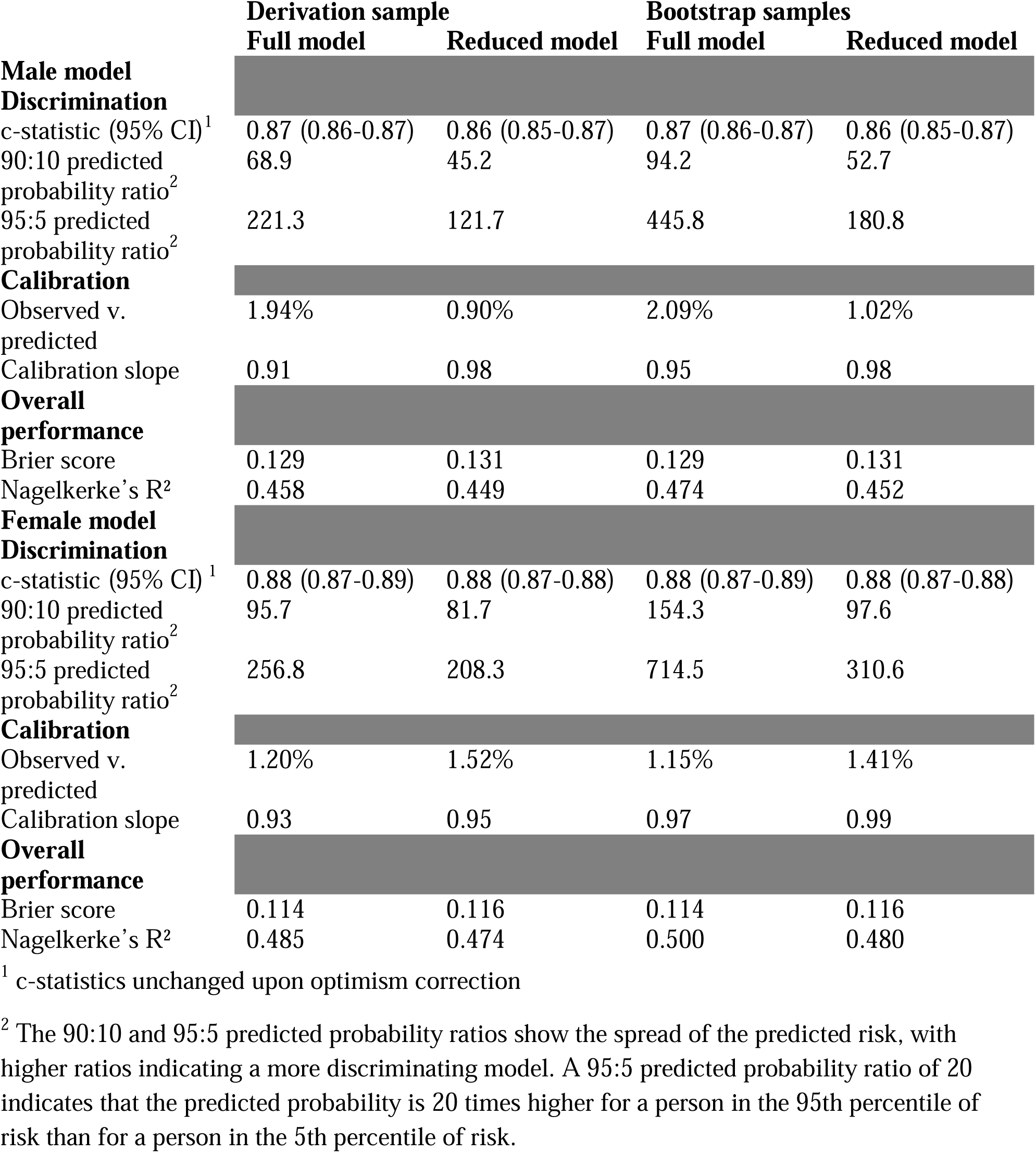
Goodness of fit summary statistics for HTNPoRT in full and reduced models across the derivation and bootstrap samples.

|  | Derivation sample |  | Bootstrap samples |  |
| --- | --- | --- | --- | --- |
|  | Full model | Reduced model | Full model | Reduced model |
| **Male model** |  |  |  |  |
| **Discrimination** |  |  |  |  |
| c-statistic (95% CI)<sup>1</sup> | 0.87 (0.86-0.87) | 0.86 (0.85-0.87) | 0.87 (0.86-0.87) | 0.86 (0.85-0.87) |
| 90:10 predicted probability ratio<sup>2</sup> | 68.9 | 45.2 | 94.2 | 52.7 |
| 95:5 predicted probability ratio<sup>2</sup> | 221.3 | 121.7 | 445.8 | 180.8 |
| **Calibration** |  |  |  |  |
| Observed v. predicted | 1.94% | 0.90% | 2.09% | 1.02% |
| Calibration slope | 0.91 | 0.98 | 0.95 | 0.98 |
| **Overall performance** |  |  |  |  |
| Brier score | 0.129 | 0.131 | 0.129 | 0.131 |
| Nagelkerke's R<sup>2</sup> | 0.458 | 0.449 | 0.474 | 0.452 |
| **Female model** |  |  |  |  |
| **Discrimination** |  |  |  |  |
| c-statistic (95% CI)<sup>1</sup> | 0.88 (0.87-0.89) | 0.88 (0.87-0.88) | 0.88 (0.87-0.89) | 0.88 (0.87-0.88) |
| 90:10 predicted probability ratio<sup>2</sup> | 95.7 | 81.7 | 154.3 | 97.6 |
| 95:5 predicted probability ratio<sup>2</sup> | 256.8 | 208.3 | 714.5 | 310.6 |
| **Calibration** |  |  |  |  |
| Observed v. predicted | 1.20% | 1.52% | 1.15% | 1.41% |
| Calibration slope | 0.93 | 0.95 | 0.97 | 0.99 |
| **Overall performance** |  |  |  |  |
| Brier score | 0.114 | 0.116 | 0.114 | 0.116 |
| Nagelkerke's R<sup>2</sup> | 0.485 | 0.474 | 0.500 | 0.480 |

<sup>1</sup> c-statistics unchanged upon optimism correction
<sup>2</sup> The 90:10 and 95:5 predicted probability ratios show the spread of the predicted risk, with higher ratios indicating a more discriminating model. A 95:5 predicted probability ratio of 20 indicates that the predicted probability is 20 times higher for a person in the 95th percentile of risk than for a person in the 5th percentile of risk.

#### Model reduction

In the reduced models, overall discriminative performance was similar in men (c-statistic: 0.86, 95% CI: 0.85-0.87) and women (c-statistic: 0.88, 95% CI: 0.87-0.88) across bootstrap samples, and the c-statistics remained unchanged after optimism correction. As seen commonly with model reduction, discrimination across predicted probability percentiles mildly degraded in men and women, but calibration slopes improved (0.98 in men and 0.99 in women across bootstrap samples). Calibration plots for the reduced models are shown in Figure 2, and additional performance metrics are presented in Table 3.

**Figure 2.**
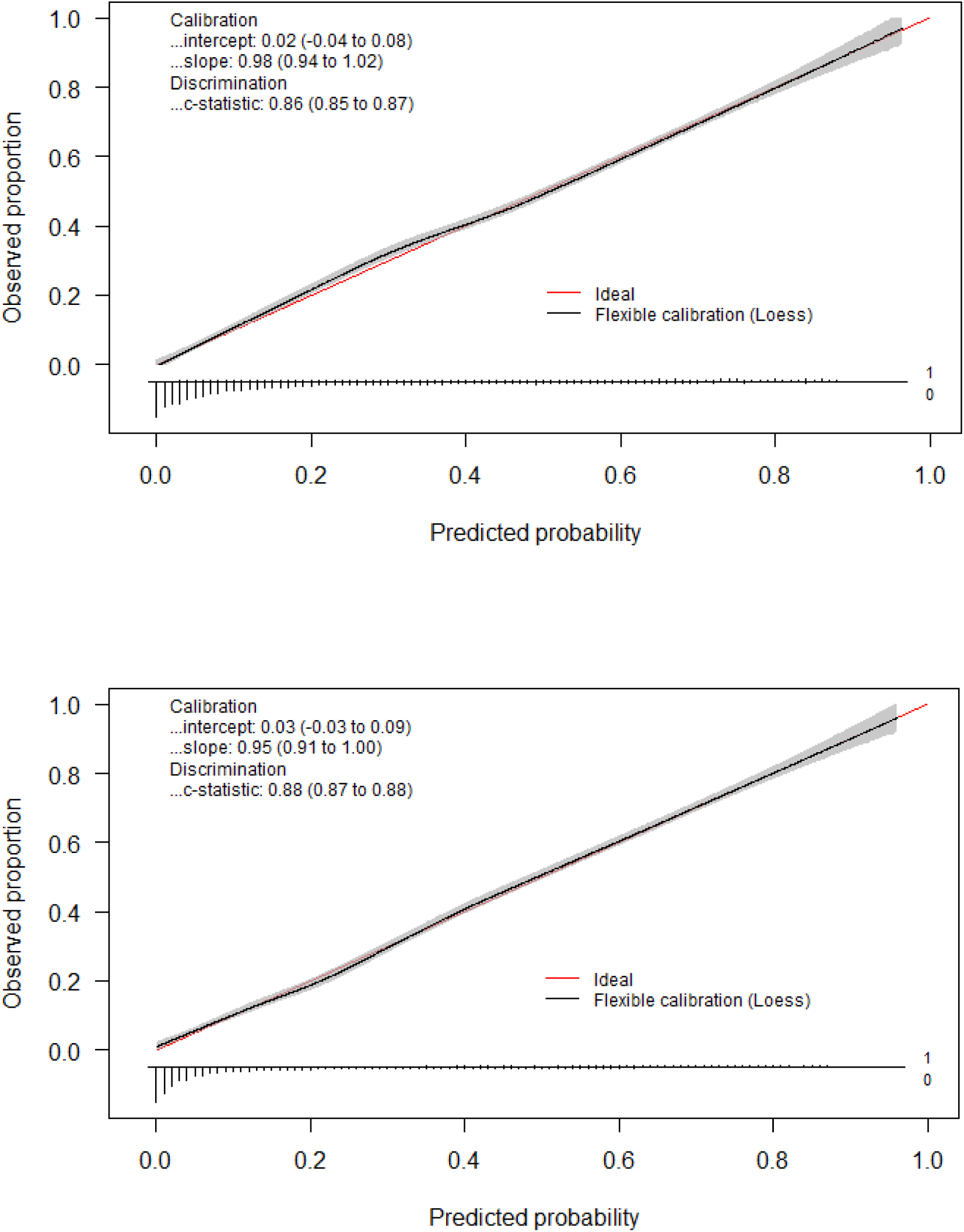
Calibration plots for reduced models; predicted hypertension probability versus observed hypertension proportion in men (top) and women (bottom)

#### Performance in policy-relevant subgroups and sensitivity analyses

The full model was well calibrated across all 181 predefined policy-relevant subgroups (stratified by sociodemographics, health status, etc.) where hypertension prevalence was at least 5% (Appendix 6). The reduced model also performed well, showing poor calibration in only two subgroups: men with chronic kidney disease and men who are heavy drinkers (Appendix 7). All model performance metrics and predicted risks remained similar and within expectations during the sensitivity analyses outlined earlier. Notably when using 130/80 mm Hg blood pressure cut-offs to define hypertension for all respondents, discrimination decreases somewhat in both men (c-statistics: 0.76-0.77) and women (c-statistics: 0.81-0.82) but is still very high considering the increased hypertension prevalence (Appendix 8).

### Model presentation

#### Example of a hypertension risk profile

Figure 3 demonstrates a risk profile for a hypothetical 74-year-old man using HTNPoRT. His predicted probability of currently having hypertension is 75.3%, substantially higher than the population average of 27.6% for men. The profile, generated using SHAP values, shows how individual factors contribute to his overall probability. His age is the most influential factor, associated with a 28.0% increase in probability relative to the average. His diabetes diagnosis and BMI of 35.3 kg/m² are associated with further increases of 20.2% and 6.9%, respectively. Conversely, his lack of a family history of hypertension is associated with a 5.6% decrease in probability. This profile illustrates that the individual’s high probability is strongly associated with his older age, diabetes, and higher BMI.

**Figure 3.**
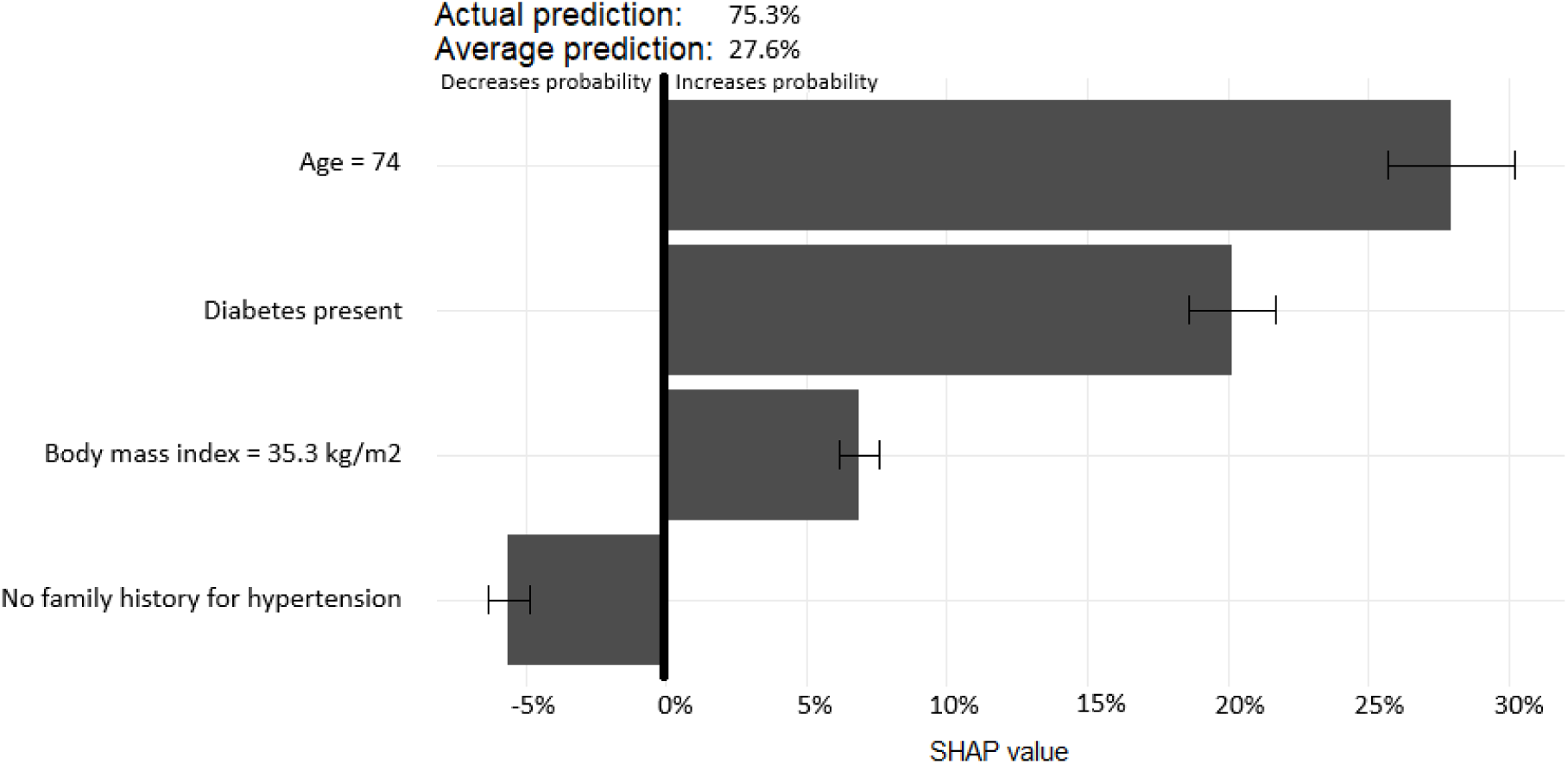
Hypertension risk profile of a 74-year-old male with a predicted hypertension probability of 75.3%.

#### Predicted hypertension risks in Canada

Table 4 shows the mean predicted hypertension risk across both the overall population and risk groups present in all four models. The average risk of hypertension was about 27% for men and 24% for women. Risks increased steeply from ages 20–25 to 75–79, more than three-fold with diabetes, and nearly double with obesity.

**Table 4.** Mean predicted probabilities across HTNPoRT study populations and risk groups from predictors present in all models.

| Variable | Group | Mean predicted probability (%) |  |  |  |
| --- | --- | --- | --- | --- | --- |
|  |  | Male full model | Female full model | Male reduced model | Female reduced model |
| **Overall** | N/A | 27.3 | 24.4 | 27.6 | 24.3 |
| **Age** |  |  |  |  |  |
|  | 20-24 years | 1.0 | 0.7 | 1.1 | 0.6 |
|  | 25-39 years | 2.0 | 1.1 | 2.2 | 1.1 |
|  | 30-34 years | 4.7 | 2.4 | 4.8 | 2.4 |
|  | 35-39 years | 8.2 | 4.9 | 8.3 | 4.8 |
|  | 40-44 years | 13.0 | 8.6 | 13.3 | 8.3 |
|  | 45-49 years | 19.7 | 15.0 | 20.2 | 15.1 |
|  | 50-54 years | 28.2 | 24.4 | 28.2 | 23.9 |
|  | 55-59 years | 38.4 | 34.1 | 38.6 | 33.7 |
|  | 60-64 years | 47.0 | 43.1 | 47.9 | 43.5 |
|  | 65-69 years | 56.3 | 53.2 | 56.4 | 53.1 |
|  | 70-74 years | 64.8 | 61.1 | 64.9 | 61.0 |
|  | 75-79 years | 71.0 | 71.4 | 71.2 | 70.9 |
| **Hypertension family history** |  |  |  |  |  |
|  | Yes | 34.3 | 31.4 | 34.9 | 31.2 |
|  | No | 20.3 | 15.9 | 20.4 | 16.0 |
| **Body mass index** |  |  |  |  |  |
|  | Underweight | 14.8 | 10.3 | 15.1 | 10.1 |
|  | Normal | 22.7 | 19.9 | 23.1 | 20.0 |
|  | Overweight | 29.5 | 29.0 | 30.0 | 29.0 |
|  | Obese | 42.2 | 38.3 | 42.2 | 38.2 |
| **Diabetes** |  |  |  |  |  |
|  | Yes | 72.9 | 69.7 | 72.6 | 69.5 |
|  | No | 21.4 | 20.3 | 21.8 | 20.2 |

## Discussion

The Hypertension Population Risk Tool (HTNPoRT) is a classification algorithm that is both discriminating and well-calibrated, having been developed and validated in a nationally representative, multiethnic Canadian population. A key feature of the model is its development using easily collected population-based survey data. For individuals, the algorithm can enhance personal risk awareness and facilitate conversations with healthcare providers about hypertension screening and prevention. People who are not screened can therefore use risk prediction models such as HTNPoRT to understand their risk of hypertension, thereby making more informed decisions about their cardiovascular health.

However, equally or more importantly, uses of HTNPoRT are for risk communication, health planning, and knowledge generation in populations. For health planners, it can help identify populations with a high burden of potential or those with undiagnosed hypertension who may benefit from targeted outreach and prevention programs (Manuel et al., 2012). Such identified populations can include sociodemographic groups with disproportionate hypertension burden. HTNPoRT can also be used to support estimations of screening yield, cost-effectiveness, and monitoring progress to reducing inequities in hypertension risk.

### Strengths and methodological considerations

The favourable performance of HTNPoRT can be attributed to the study’s strengths. First, the use of the CHMS provided a large, representative sample, which afforded the statistical power to develop a highly specified model with flexible non-linear terms and interactions. While increased model complexity can increase the risk of overfitting, this was mitigated by pre-specifying all predictors. Including a medication inventory in the hypertension definition is a strength, as it ensures the model captures individuals with both controlled and uncontrolled hypertension, reflecting the true population burden.

A third strength is the model’s ability to describe the relative importance of key hypertension predictors in the presence of interactions and non-linear terms. As expected, age was the most powerful predictor of hypertension status. Among modifiable factors, measures of adiposity were among the most important contributors to risk. While there is debate about the clinical limitations of body mass index (BMI), our analysis confirmed that it is a simple, practical, and highly predictive component for this tool. Our analysis showed a sex-specific nuance: while the waist-to-height ratio was very collinear with BMI, it was slightly more strongly associated with hypertension in men (see note 3 of Table 1). It did not improve the overall model discrimination or calibration when substituted, possibly reflecting known sex differences in fat distribution (Chang et al., 2018; Salim et al., 2021). Consequently, BMI was retained as the sole adiposity measure in accordance with our prespecified protocol. This decision prioritizes utility without sacrificing accuracy; height and weight are more frequently known and self-reported by the public than waist circumference, minimizing barriers to tool use.

### Study in context

A recent review of 75 hypertension prediction models provides context for HTNPoRT’s performance (Chowdhury et al., 2022). The review reported a pooled c-statistic of 0.75 (95% CI: 0.73-0.77), with a trade-off: complex data, such as genetics or biomarkers, achieved high discrimination (c-statistic > 0.80), while the only comparable model using readily available data performed modestly (c-statistic: 0.74) (Wang et al., 2015). HTNPoRT bridges this gap, achieving discriminative ability (c-statistic: 0.86-0.88) comparable to models that rely on complex clinical data, yet using only readily available data. This favourable performance is likely attributable to its highly specified modelling approach, which flexibly accounted for key non-linear relationships and interactions. Meanwhile, our study showed that hypertension risk among Canadians increases remarkably with older age, diabetes, and obesity, building on findings from an earlier study (Leung et al., 2019).

### Limitations

This study has several limitations. First, the predictors were restricted to those available in the CHMS, and other unmeasured factors may also contribute to hypertension risk. Second, while the CHMS is nationally representative, it excludes certain populations, such as on-reserve Indigenous peoples, which may limit the model’s generalizability to these specific groups. Finally, reliance on self-reported predictors introduces a risk of measurement error; however, the model performed well despite this potential source of error.

The issue of measurement error is particularly relevant for the diet predictor. A notable finding was the low predictive influence of diet in our model, which contrasts with the strong causal evidence from intervention trials of the DASH diet (Mills et al., 2020). This discrepancy likely highlights two key issues: the difficulty of capturing complex dietary patterns with survey questions, and the high degree of collinearity between diet and other predictors, such as adiposity. Therefore, while HTNPoRT is an effective risk stratification tool, it is not designed to estimate the potential benefits of specific dietary interventions, and any associations between risk factors and hypertension should not be causally interpreted.

A key consideration is the model’s generalizability to other countries. The model’s performance across different healthcare systems and in countries with varying demographic profiles and baseline hypertension risks is unknown. The transportability of the model’s effects may also vary by predictor. For instance, the strong influence of a universal biological factor, such as adiposity, is likely to be robust across settings (O’Donnell et al., 2016; Yusuf et al., 2004). In contrast, the effect size for age may be more context-dependent, being a proxy for cumulative exposures differing between populations. Fortunately, this challenge can be readily addressed, as the age term is particularly amenable to recalibration using local, age-stratified hypertension prevalence data from a new target population. HTNPoRT was developed to support transportability across populations by centring all predictors and interactions, as well as reporting predictor correlations and other data (see analytical code).

Despite these limitations, HTNPoRT was designed with several strengths that support its robustness and potential for broad application. It was developed in a diverse, multiethnic population, enhancing its relevance for varied community settings. The use of a transparent logistic regression framework and techniques like predictor centering - which is specifically intended to facilitate recalibration in new populations - provide a clear and adaptable foundation for future external validation studies.

## Conclusions

This study demonstrates that a high-performing and interpretable predictive model for hypertension can be developed using only readily available data from population-based health surveys. The resulting algorithm, HTNPoRT, achieves discriminative performance comparable to that of models that rely on complex clinical data. There are several next steps to support HTNPoRT use or the use of other models that use the HTNPoRT approach. First, external validation in other countries is feasible, as the CHMS methodology is like many international health surveys. Second, future implementation science research can assess how this readily accessible risk assessment tool can best address existing needs in both clinical communication and public health planning.

## Data Availability

Canadian Health Measures Survey (CHMS) data is not available to the public. See Appendix 2.

https://osf.io/mjd7n/

https://github.com/Big-Life-Lab/htnport

## Contributions to knowledge

### What does this study add to existing knowledge?

- The Hypertension Population Risk Tool (HTNPoRT) is a diagnostic algorithm developed entirely from readily available, population-based survey data.
- Hypertension risk varies across Canada and is concentrated among high-risk individuals. (e.g., older adults, people with diabetes).

### What are the key implications for public health interventions, practice, or policy?

- Many hypertension prediction models are too complex for public health use. HTNPoRT provides a practical, generalizable tool for population-level risk assessment that achieves discrimination comparable to complex clinical models and is well-calibrated across diverse, policy-relevant subgroups, filling a critical gap in preventative health tools.
- HTNPoRT can enhance public awareness of hypertension risk and facilitate patient-provider conversations about screening and evidence-based prevention strategies.

## Notes

### Competing Interest Statement

The authors have declared no competing interest.

### Clinical Protocols

https://doi.org/10.17605/OSF.IO/V27MW

### Funding Statement

This study was funded by the grant BHI 23-111 from uOttawa Brain-Heart Interconnectome (BHI), a Canada First Research Excellence Fund (CFREF) program.

### Author Declarations

The Ottawa Health Science Network Research Ethics Board of the Ottawa Hospital Research Institute gave approval for this work.

### Summary of Updates

Title, author, and copyright information updated; Table 4 added; Supplemental files updated

