## Appendices 1-8 for "Development and Validation of the Hypertension Population Risk Tool: A Population-Based Diagnostic Algorithm for Canadians"

### **Appendix 1 – Details on deviations from registered protocol**

We adhered to the protocol with the following exceptions:

- The definition of alcohol consumption was changed from never, former, light, or moderate-to-heavy drinkers to never, low-risk (i.e., former or light), moderate, or heavy drinkers to correctly align with Canada's current Low-Risk Drinking Guideline.
- The variance inflation factor threshold for significant collinearity was lowered from 10 to 2.5 to ensure the model’s predictors were sufficiently independent from one another.
- Bivariable analysis, likelihood ratio tests, partial effects plotting, and odds ratios presentations were deemed redundant and not performed.
- Sensitivity analyses were performed to assess if model performance was affected by any missing data, other imputed datasets generated with the same MICE imputation model, skewness, possible collinear predictors, linear interactions, unadjusted blood pressures, potential anti-hypertensive medication misclassification, and new 130/80 mm Hg blood pressure cut-offs for hypertension.
- Optimism was quantified for c-statistics as a part of internal bootstrap validation.
- Final model determination was based on both model performance and reducing user burdens to ensure the final tool is robust and practical for end-users.
- Mean predicted probabilities were displayed for the overall population and select risk groups from predictors present in all full and reduced models.

### **Appendix 2 – Details on data access**

Authorized investigation team members were only allowed to have access if they were a deemed employee of Statistics Canada with a keycard to the Research Data Centre (RDC) and a login to their assigned hard drive. A keycard and login were granted after obtaining a “Reality Status” security clearance from Statistics Canada, signing a Microdata Research Contract, and affirming an Oath of Office and Secrecy to Statistics Canada. Such members were only allowed to access the study data for the proposed analyses only and were not allowed to alter or share the original data whatsoever. Any information identifying respondents in the CHMS was kept separate from the data files and can only be accessed by Statistics Canada employees involved in the collection and generation of data (i.e., no one in the investigation team). The CHMS data files available for use to derive average moderate-to-vigorous physical activity (MVPA) minutes per week contain respondents with only 4 or more valid days of accelerometer data. From the complete CHMS accelerometer files stored at Statistics Canada, an approved Statistics Canada employee identified respondents in all cycles who had 1 to 3 valid days of accelerometer data, derived their average MVPA minutes per week, and provided these values in a data file which was placed in the investigation team’s assigned hard drive and then merged to the rest of the study data for a more complete analysis. Only analyses output (not any study data) was transferred by employees at the RDC after being vetted for small cell sizes prior to release. Such output was transferred to a folder which can only by accessed on Statistics Canada’s secure network by members of the investigation team at the Health Analysis and Modelling Division. All study records will be retained securely for 10 years upon the completion of this study and then destroyed as per the requirements of the Ottawa Hospital Research Institute.

### **Appendix 3 – Details on variable definitions and derivations**

*Blood pressure.* Systolic blood pressure (SBP) and diastolic blood pressure (DBP) were both measured at the MEC with the BpTRU™ BPM-300 device, which is an automated oscillometric device that has been validated and recommended for use by Hypertension Canada. The device was administered in quiet, temperature-controlled rooms with the lights always on. Following a five-minute rest period (in which participants were asked to sit quietly, relax and refrain from moving or talking), six measurements were taken at one-minute intervals for each participant in the absence of any survey personnel, thereby reducing the risk of observer-participant interactions which can influence results.

Afterwards, the last five measurements were averaged together to determine the average SBP and DBP levels, and the average blood pressure readings were then adjusted using the following correction factors to approximate manual sphygmomanometer readings, as is standard for CHMS analyses estimating hypertension prevalence: adjusted SBP = 11.4 + (0.93 * SBP) and adjusted DBP = 15.6 + (0.83 * DBP). Such approach to ascertain blood pressure is accepted as a gold standard method for population surveys and produces reliable estimates of blood pressure for the Canadian population, without the need for highly trained staff to use stethoscopes while also reducing human errors in reading and any unwanted observer-participant interactions.

*Sociodemographic measures.* Sex at birth was defined as “male” or “female”. Age was defined as years at MEC visit. Highest education level was classified as “less than secondary school graduation”, “secondary school graduation”, or “post-secondary graduation”. Marital status was defined as “married or common-law”, “widowed, separated, or divorced”, or “single and never married”. Working status was based on respondents’ answers to the question of whether they were employed as of the previous week.

*Psychosocial measures.* Psychosocial measures included respondents’ self-rated mental health, self-perceived stress, and sense of belonging to the community, each encoded on ordinal scales.

*Health status measures.* Family history for hypertension was defined as if respondents ever had an immediate family member with hypertension (excluding during pregnancy). Body mass index was measured in kg/m^2^ upon dividing weight by height squared, while waist-to-height ratio was displayed as a percentage upon dividing waist circumference by height.

*Health behaviours.* Alcohol consumption was defined as whether respondents were either never, low-risk (i.e., former or light), moderate, or heavy drinkers according to Canada's Low-Risk Alcohol Drinking Guideline. Smoking was defined as whether respondents were current smokers, former smokers, or never smokers. Average moderate-to-vigorous physical activity (MVPA) minutes per week were computed from week-long accelerometer data containing at least one day of valid data (see Appendix 2 for further details). Fruit and vegetable consumption was derived as the sum of frequency of daily consumption of select fruit, fruit juices, tomatoes, potatoes, greens, collards, and others. Sleep duration was defined as hours of sleep per night.

*Chronic conditions.* Diabetes was defined as having a positive self-report, having a level of serum glycated hemoglobin A1c of 6.5% or higher, or having taken a glucose-lowering medication in the month before the survey. Chronic kidney disease was defined as having an estimated glomerular filtration rate (derived from serum creatine, age, sex, and ethnicity) less than 60 mL/min/1.73 m^3^.

*Medication use.* Current medications were recorded during the household and clinic interviews, and these were assigned to codes from the Anatomical Therapeutic Chemical (ATC) classification system, corresponding to beta blockers, agents acting on the renin-angiotensin system, thiazide diuretics, calcium channel antagonists, other antihypertensive agents, as well as glucose-lowering medications.

### **Appendix 4 – Unimputed and unweighted characteristics of the study populations for HTNPoRT**

|  | **Male^1^** | **Female^1^** |
| --- | --- | --- |
| **Overall** |  | |
| Total | 9,633 | 10,010 |
| Hypertensive | 2,681 (27.8%) | 2,471 (24.7%) |
| – Uncontrolled | 1,300 (13.5%) | 1,238 (12.4%) |
| – Controlled | 1,381 (14.3%) | 1,233 (12.3%) |
| **Sociodemographic measures** |  | |
| Age (years) | 47 (36, 62) | 47 (36, 63) |
| Marital status |  | |
| – Married or common-law | 6,900 (71.6%) | 6,064 (60.6%) |
| – Widowed, separated, or divorced | 1,083 (11.2%) | 2,211 (22.1%) |
| – Single and never married | 1,646 (17.1%) | 1,728 (17.3%) |
| – Missing | 4 (0.04%) | 7 (0.07%) |
| Highest education level |  | |
| – Less than secondary school graduation | 1,122 (11.7%) | 1,140 (11.4%) |
| – Secondary school graduation | 1,691 (17.6%) | 1,794 (17.9%) |
| – Post-secondary school graduation | 6,714 (69.7%) | 6,996 (69.9%) |
| – Missing | 106 (1.1%) | 80 (0.8%) |
| Working status |  | |
| – Has a job | 6,816 (70.8%) | 5,892 (58.9%) |
| – Does not have a job | 2,772 (28.8%) | 4,082 (40.8%) |
| – Missing | 45 (0.5%) | 36 (0.4%) |
| **Psychological measures** |  | |
| Self-rated mental health |  | |
| – Poor or fair | 566 (5.9%) | 785 (7.8%) |
| – Good, very good, or excellent | 9,036 (93.8%) | 9,190 (91.8%) |
| – Missing | 31 (0.3%) | 35 (0.4%) |
| Self-perceived stress |  | |
| – Not at all to a bit | 7,614 (79%) | 7,592 (75.8%) |
| – Quite a bit or extremely | 2,015 (20.9%) | 2,415 (24.1%) |
| – Missing | 4 (0.04%) | 3 (0.03%) |
| Sense of belonging |  | |
| – Strong | 6,358 (66%) | 6,910 (69%) |
| – Weak | 3,226 (33.5%) | 3,048 (30.5%) |
| – Missing | 49 (0.5%) | 52 (0.5%) |
| **Health status measures** |  | |
| Hypertension family history |  | |
| – Yes | 2,839 (29.5%) | 3,432 (34.3%) |
| – No | 3,155 (32.8%) | 3,090 (30.9%) |
| – Missing | 486 (5%) | 400 (4%) |
| – Not asked | 3,153 (32.7%) | 3,088 (30.9%) |
| Body mass index (kg/m^2^) | 27.2 (24.6, 30.4) | 26.2 (22.8, 30.8) |
| – Missing | 53 (0.6%) | 68 (0.7%) |
| **Health behaviours** |  | |
| Alcohol consumption |  | |
| – Never drank | 913 (9.5%) | 1,776 (17.7%) |
| – Low-risk drinker | 7,547 (78.4%) | 7,506 (75%) |
| – Moderate drinker | 426 (4.4%) | 406 (4.1%) |
| – Heavy drinker | 645 (6.7%) | 265 (2.6%) |
| – Missing | 102 (1.1%) | 57 (0.6%) |
| Smoking status |  | |
| – Current smoker | 1,943 (20.2%) | 1,656 (16.5%) |
| – Former smoker | 3,314 (34.4%) | 2,872 (28.7%) |
| – Never smoker | 4,340 (45.1%) | 5,459 (54.5%) |
| – Missing | 36 (0.4%) | 23 (0.2%) |
| Physical activity minutes (minutes/week) | 103 (39, 207) | 79 (24, 166) |
| – Missing | 977 (10.1%) | 959 (9.6%) |
| Daily fruit and vegetable consumption (times consumed/day) | 3.03 (2.14, 4.19) | 3.63 (2.61, 4.86) |
| Sleep duration (hours/night) | 7.00 (6.00, 8.00) | 7.00 (6.50, 8.00) |
| – Missing | 4 (0.04%) | 4 (0.04%) |
| – Not asked | 3,153 (32.7%) | 3,088 (30.9%) |
| **Chronic conditions** |  | |
| Diabetes |  | |
| – Yes | 1,021 (10.6%) | 790 (7.9%) |
| – No | 7,658 (79.5%) | 8,549 (85.4%) |
| – Missing | 954 (9.9%) | 671 (6.7%) |
| Chronic kidney disease |  | |
| – Yes | 511 (5.3%) | 710 (7.1%) |
| – No | 8,982 (93.2%) | 9,107 (91%) |
| – Missing | 140 (1.5%) | 193 (1.9%) |

^1^ Median (Q1, Q3); n (unweighted counts) (unweighted %)

### **Appendix 5 – Beta coefficients of full and reduced models**

| **Variable** | **Male** | | **Female** | |
| --- | --- | --- | --- | --- |
|  | **Full model** | **Reduced model** | **Full model** | **Reduced model** |
| Intercept | -2.03938833 | -1.915239452 | -2.327440614 | -2.248472641 |
| First RCS (restricted cubic spline) component for age (Age) | 0.831388368 | 0.59633291 | 0.003859081 | 0.227008328 |
| Second RCS component for age (Age') | -1.266023104 | -0.983341842 | 0.179718757 | -0.716984247 |
| Third RCS component for age (Age'') | 2.435884081 | 2.096381011 | -0.130706805 | 2.081296015 |
| Widowed, separated, or divorced | 0.011753056 |  | 0.019019769 |  |
| Single and never married | 0.039808873 |  | 0.101933808 |  |
| Secondary school graduate only | 0.037040512 |  | 0.142132526 |  |
| Never finished secondary school | 0.346332376 |  | 0.106132074 |  |
| Working status | -0.13466716 |  | 0.100371978 |  |
| Self-rated mental health | -0.077297363 |  | -0.165090454 |  |
| Self-perceived stress | -0.068451241 |  | -0.024455701 |  |
| Sense of belonging | 2.416491771 |  | 0.836853622 |  |
| Hypertension family history | 0.811156601 | 0.782228096 | 0.892403039 | 0.85999173 |
| First RCS component for body mass index (BMI) | 0.870223719 | 0.79711022 | 0.225767213 | 0.145969454 |
| Second RCS component for body mass index (BMI') | -0.844962018 | -0.802155451 | 0.015627609 | 0.129755466 |
| Low-risk (former or light) drinker | 0.18512661 |  | -0.230831958 |  |
| Moderate drinker | 0.601096708 |  | 0.306115634 |  |
| Heavy drinker | 1.25731058 |  | 0.368089047 |  |
| First RCS component for physical activity minutes (Exercise) | -0.008612277 |  | 0.016143898 |  |
| Second RCS component for physical activity minutes (Exercise') | 0.013763409 |  | -0.035752916 |  |
| Former smoker | 1.045587243 |  | 1.819165382 |  |
| Current smoker | -1.118775738 |  | 0.920389654 |  |
| Sleep duration | 0.763936189 |  | -1.331753838 |  |
| Daily fruit and vegetable consumption | -0.00108793 |  | -0.024468502 |  |
| Diabetes | 6.03482037 | 5.79643251723617 | 1.066921003 | 2.64266719888541 |
| Chronic kidney disease | 7.628826031 |  | 5.106698493 |  |
| Age:Sense of belonging | -0.054972746 |  | -0.015929847 |  |
| Age':Sense of belonging | 0.101922279 |  | 0.013528891 |  |
| Age'':Sense of belonging | -0.238999574 |  | 0.000928856 |  |
| Age:BMI | -0.02139886 | -0.018665302 | -0.004544447 | -0.001968393 |
| Age':BMI | 0.045712387 | 0.036399545 | 0.027980957 | 0.021593308 |
| Age'':BMI | -0.101684598 | -0.07722547 | -0.083259879 | -0.068405591 |
| Age:BMI' | 0.023698803 | 0.021531623 | -0.000706525 | -0.004475157 |
| Age':BMI' | -0.050398253 | -0.040003965 | -0.014938772 | -0.003069774 |
| Age'':BMI' | 0.103656052 | 0.07560878 | 0.055094881 | 0.026362919 |
| Age:Exercise | 0.000353785 |  | -0.000447063 |  |
| Age':Exercise | -0.002557437 |  | 0.000207842 |  |
| Age'':Exercise | 0.008213978 |  | 0.000467554 |  |
| Age:Exercise' | -0.000556 |  | 0.000943675 |  |
| Age':Exercise' | 0.0036823 |  | -0.000968508 |  |
| Age'':Exercise' | -0.01185042 |  | 0.000820402 |  |
| Age:Former smoker | -0.017572272 |  | -0.051530597 |  |
| Age':Former smoker | -0.062937645 |  | 0.129882254 |  |
| Age'':Former smoker | 0.261326278 |  | -0.274679026 |  |
| Age:Current smoker | 0.029196794 |  | -0.033225161 |  |
| Age':Current smoker | -0.108598324 |  | 0.13967307 |  |
| Age'':Current smoker | 0.351105878 |  | -0.417416334 |  |
| Age:Sleep | -0.019244631 |  | 0.045685775 |  |
| Age':Sleep | 0.023972894 |  | -0.15311149 |  |
| Age'':Sleep | -0.032741755 |  | 0.360599236 |  |
| Age:Diabetes | -0.093363163 | -0.093051063 | 0.017223791 | -0.024490926 |
| Age':Diabetes | 0.103215272 | 0.120165708 | 0.003858444 | 0.102039921 |
| Age'':Diabetes | -0.260382646 | -0.304303246 | -0.158014951 | -0.366964525 |
| Age:Chronic kidney disease | -0.18195042 |  | -0.087906113 |  |
| Age':Chronic kidney disease | 0.386112756 |  | 0.054376039 |  |
| Age'':Chronic kidney disease | -0.853806739 |  | 0.002674929 |  |

### **Appendix 6 – Calibration across policy-relevant subgroups for full models**

**Male full model:**

**X – excluded subgroup with observed estimate less than 5%**

*** – subgroup with difference between observed and predicted estimates over 20%**

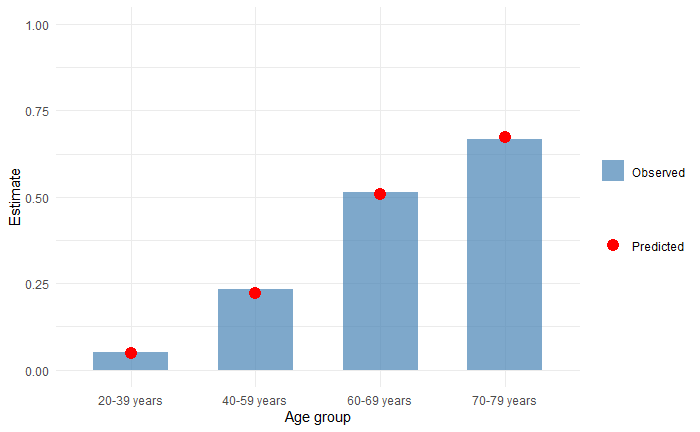

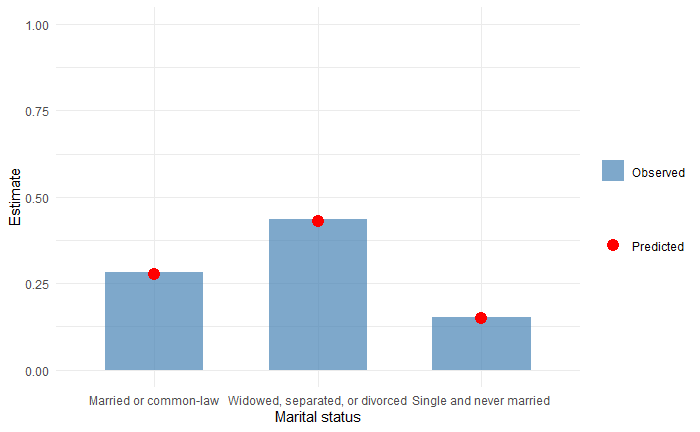

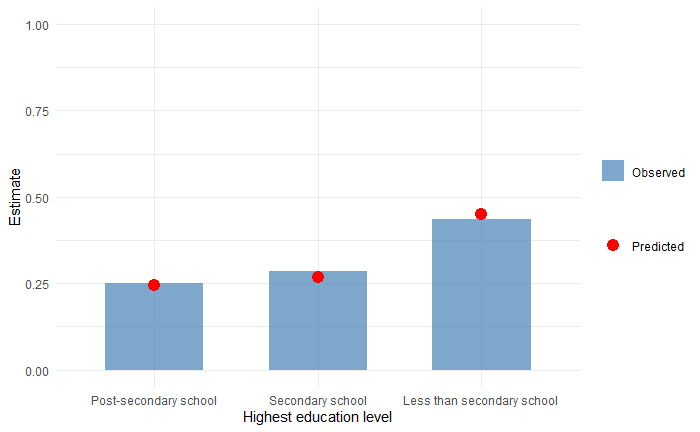

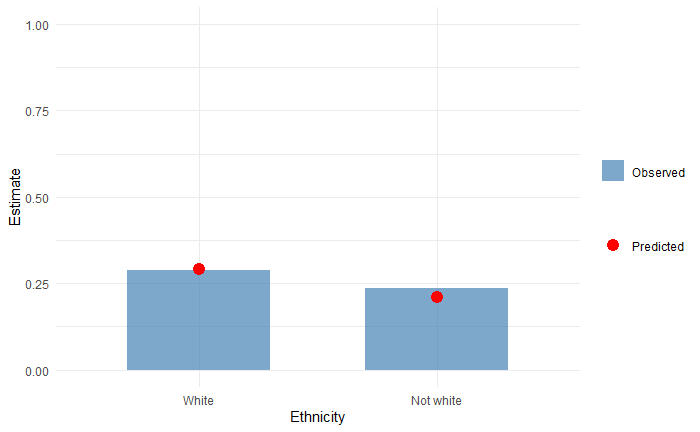

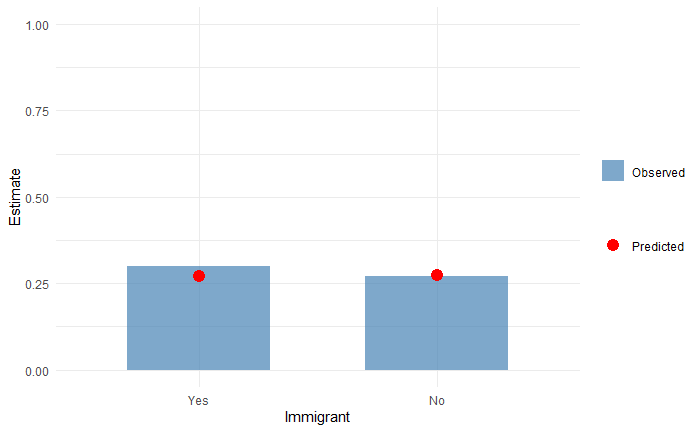

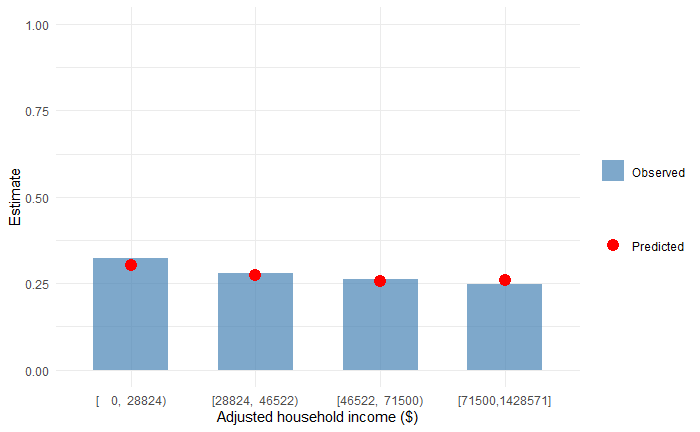

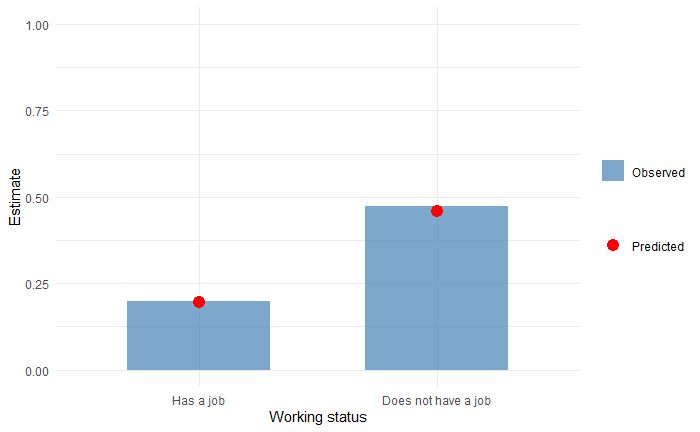

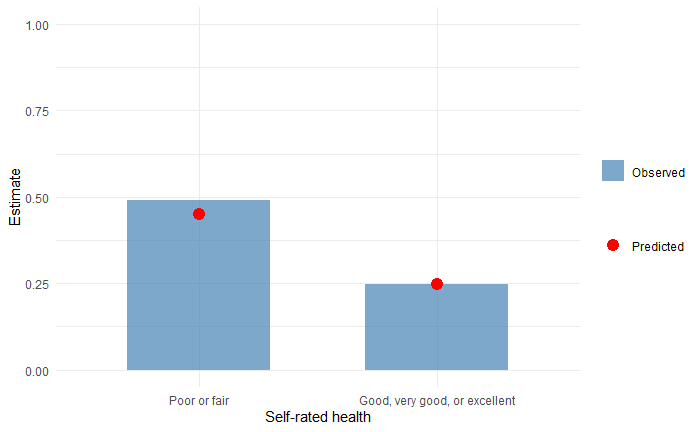

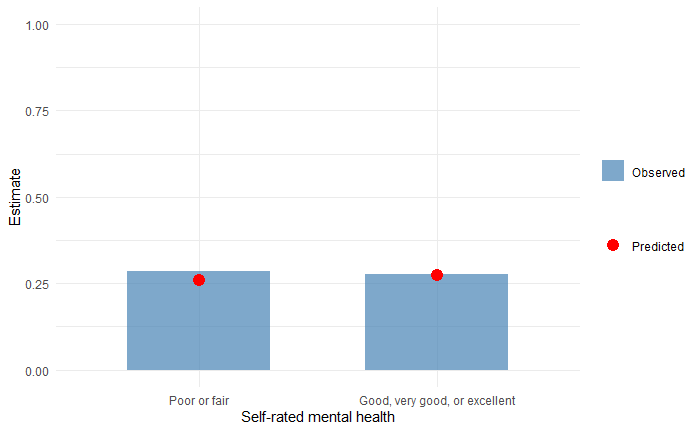

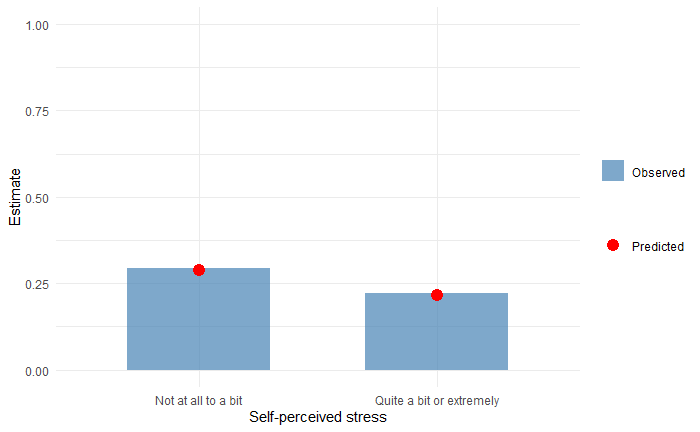

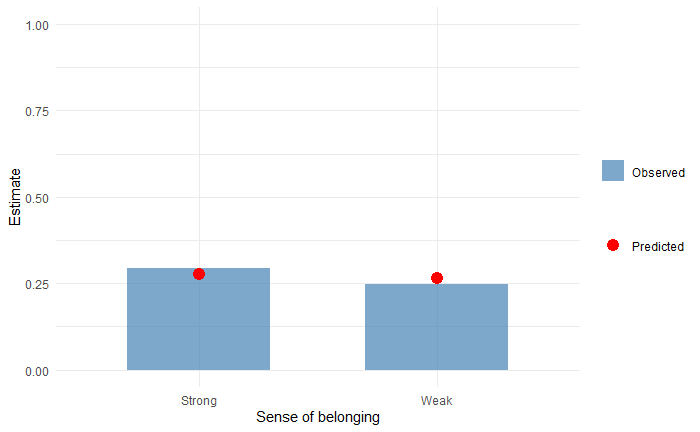

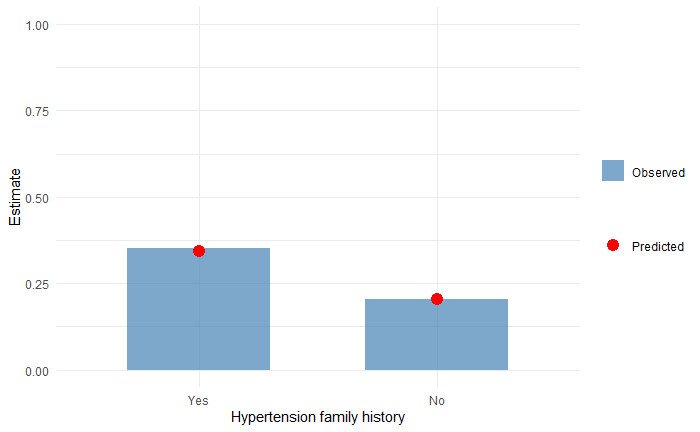

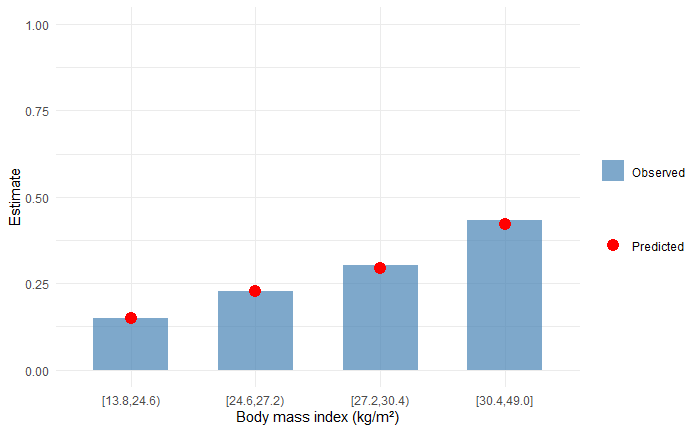

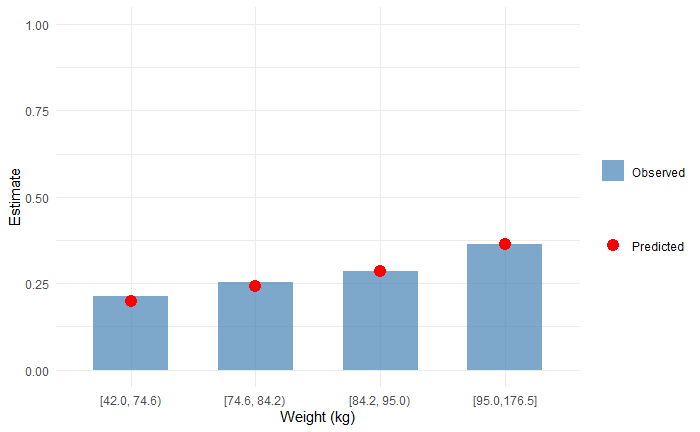

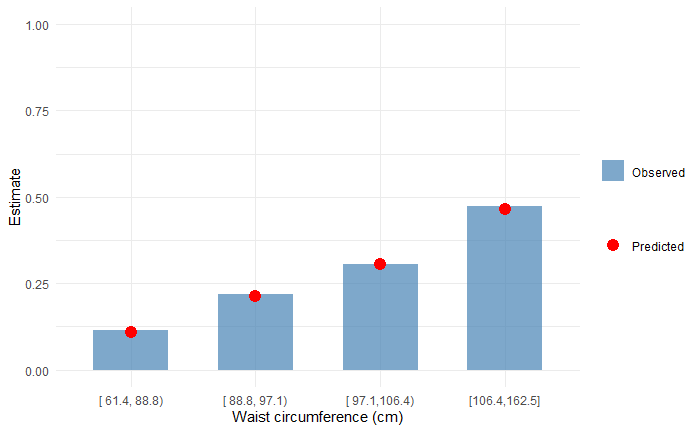

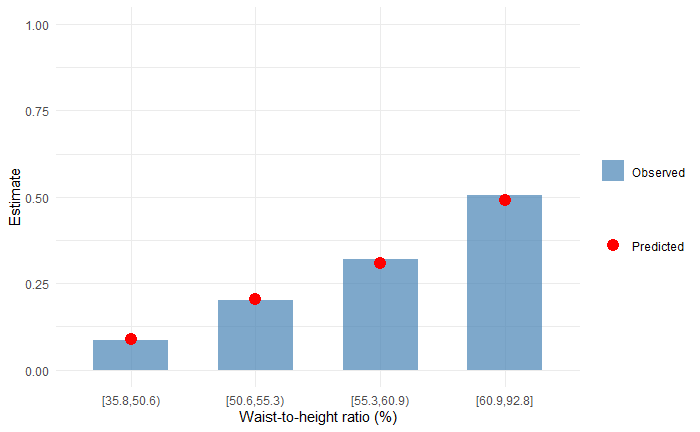

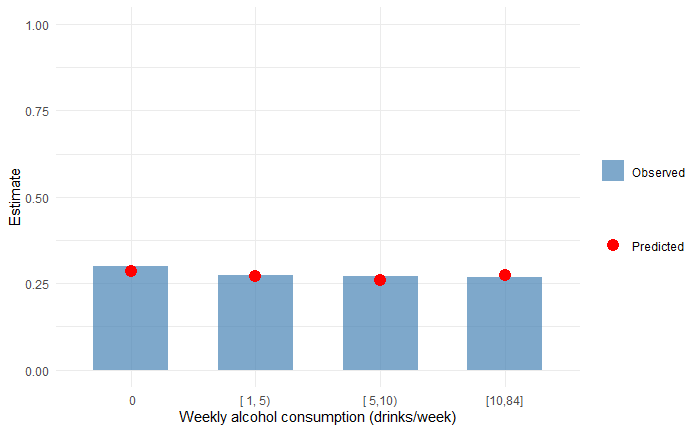

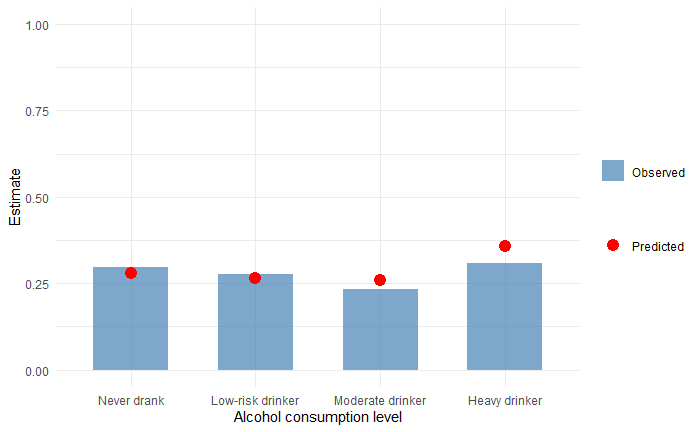

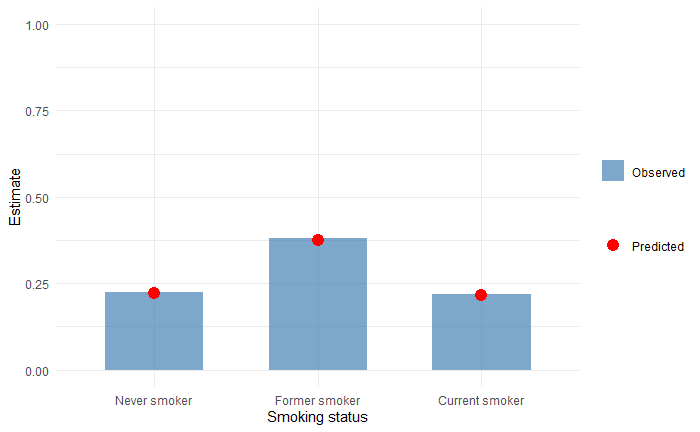

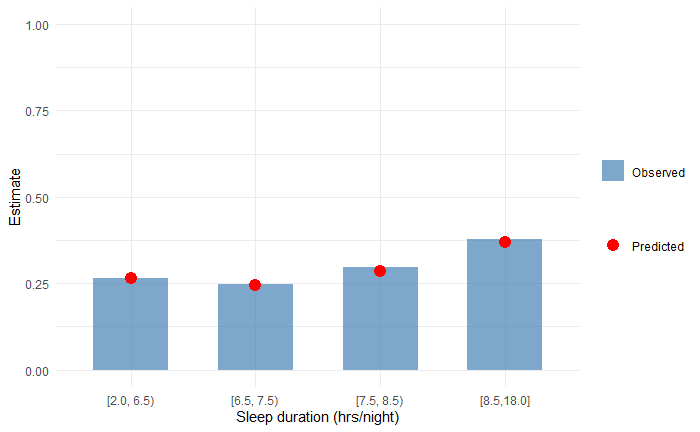

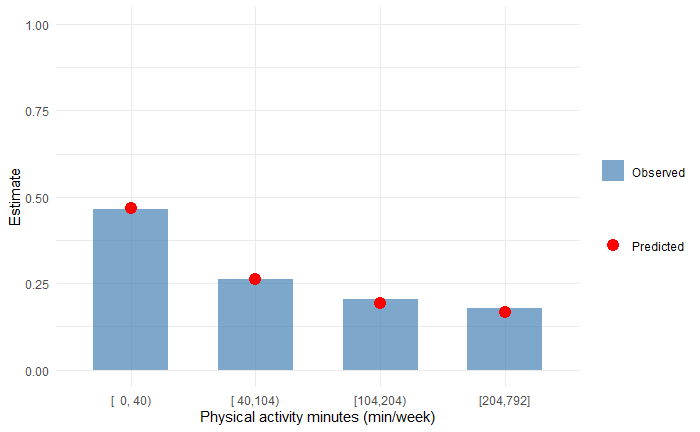

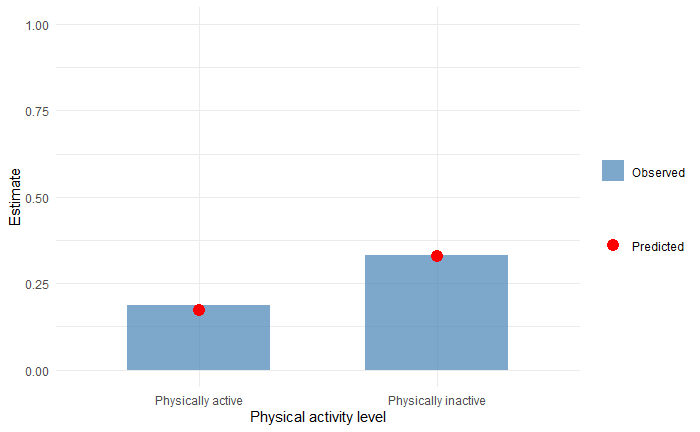

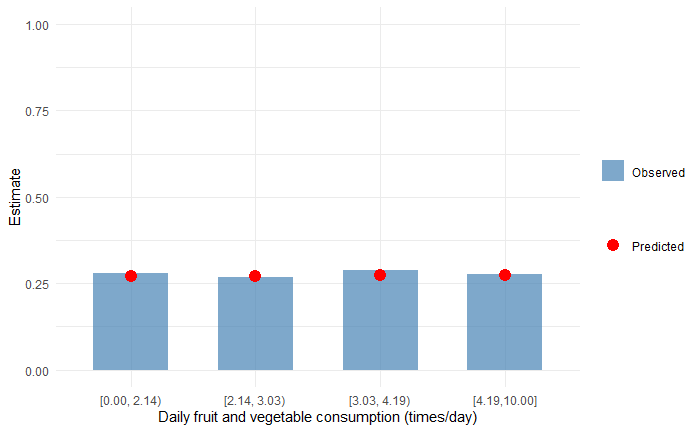

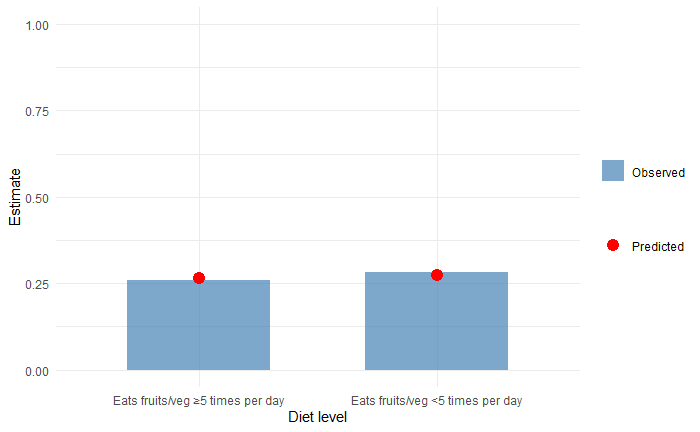

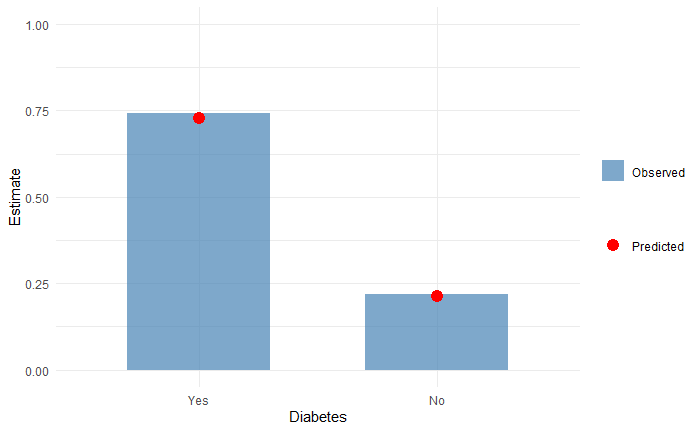

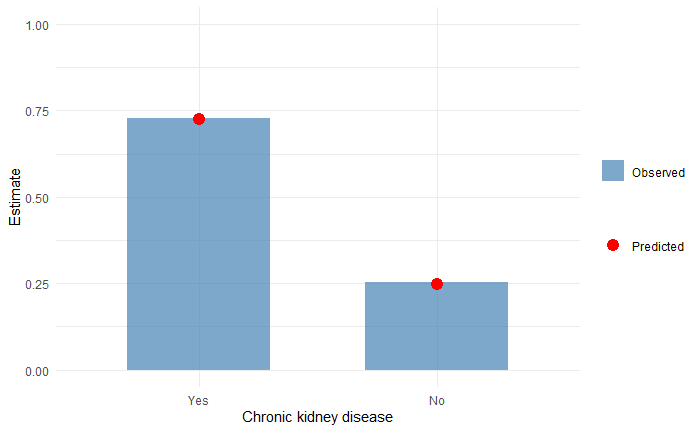

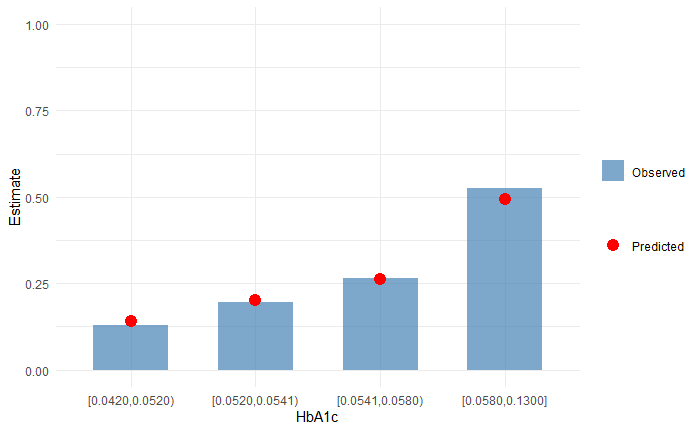

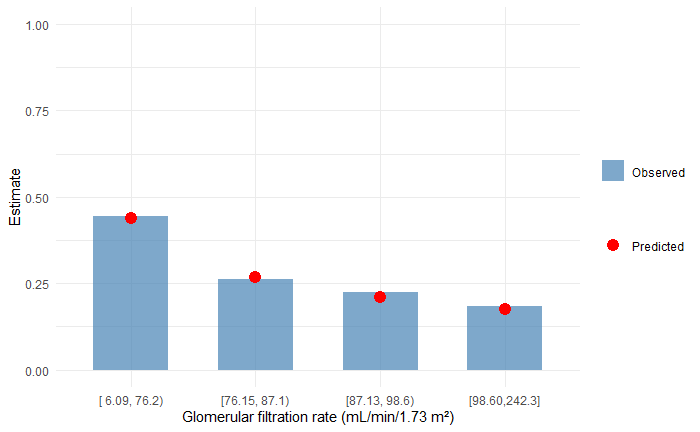

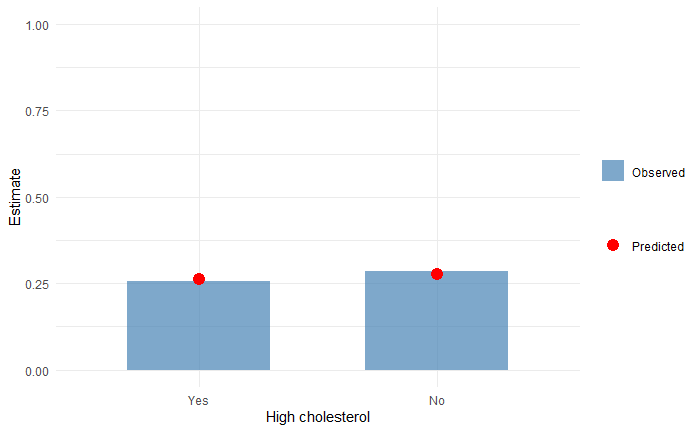

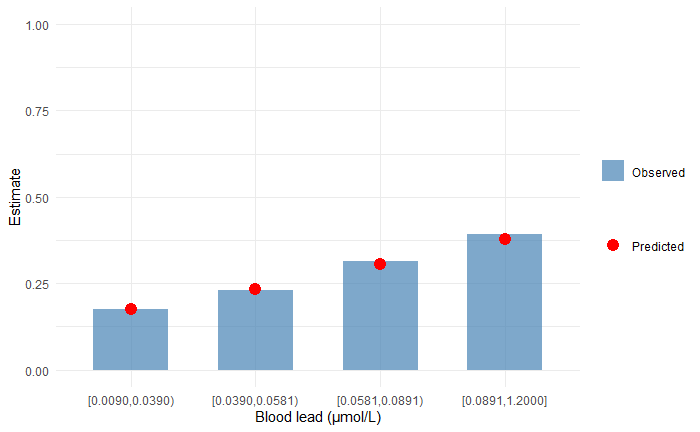

**Female full model:**

**X – excluded subgroup with observed estimate less than 5%**

*** – subgroup with difference between observed and predicted estimates over 20%**

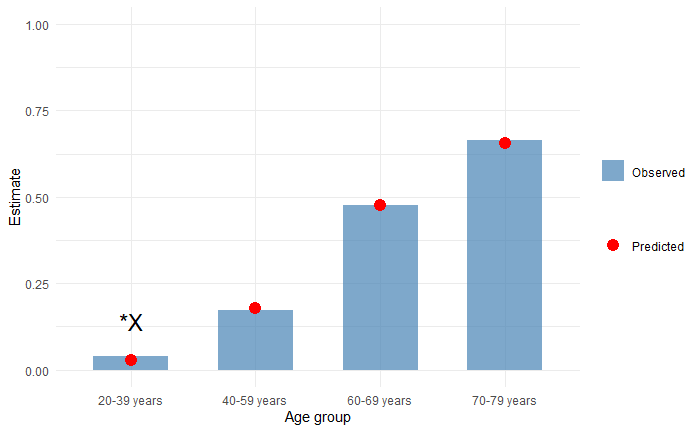

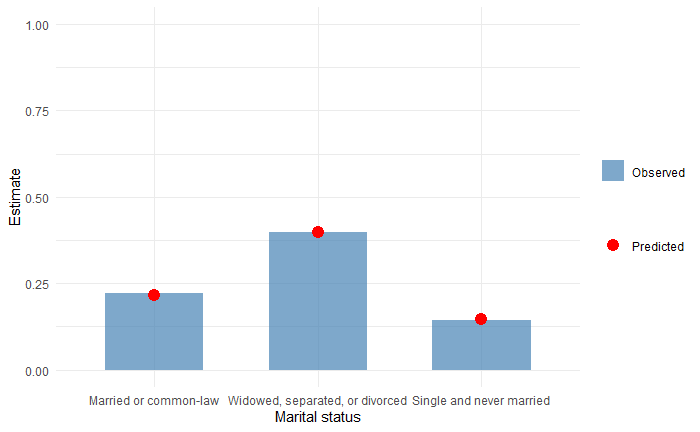

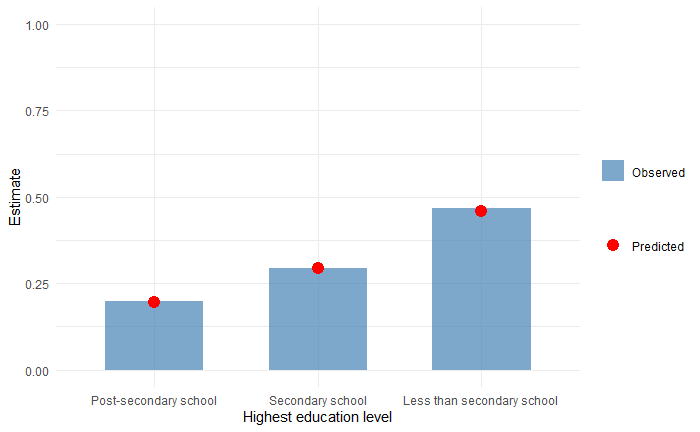

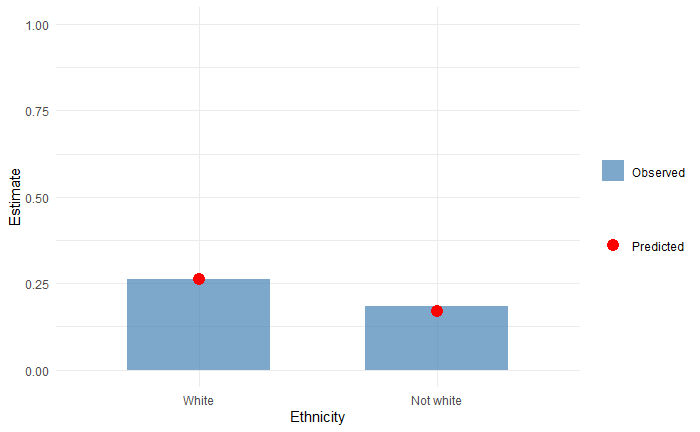

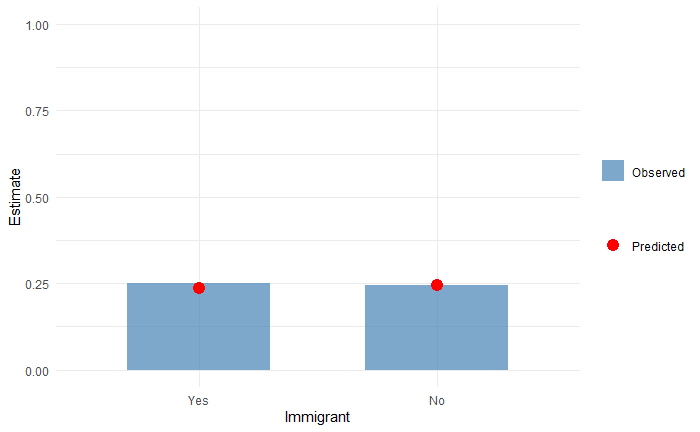

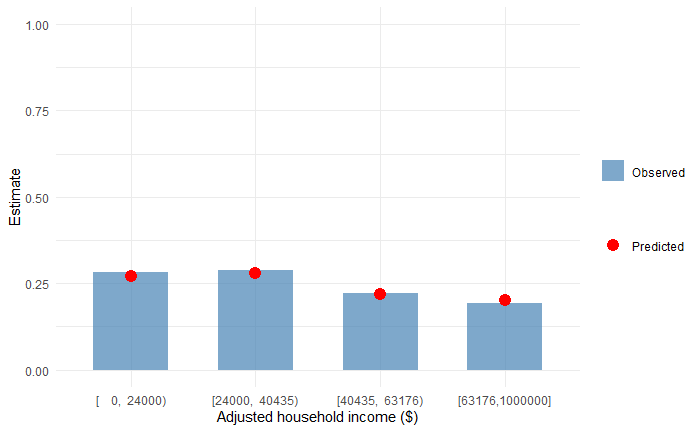

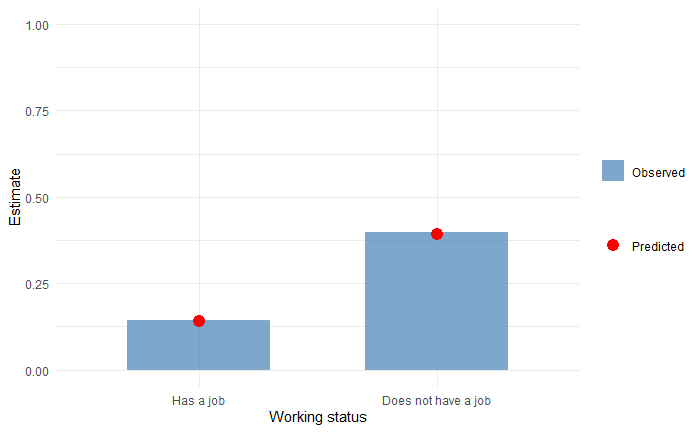

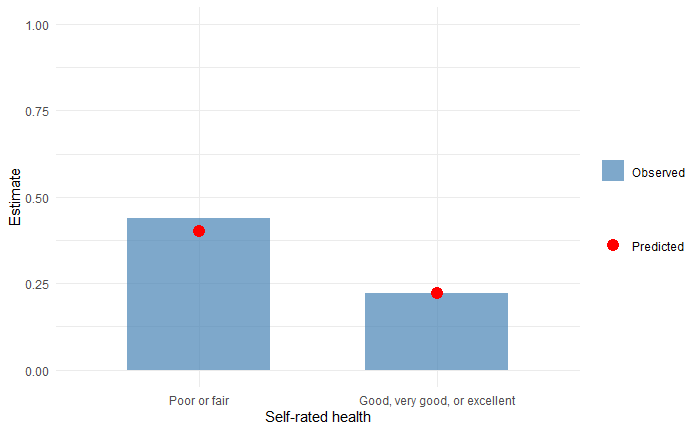

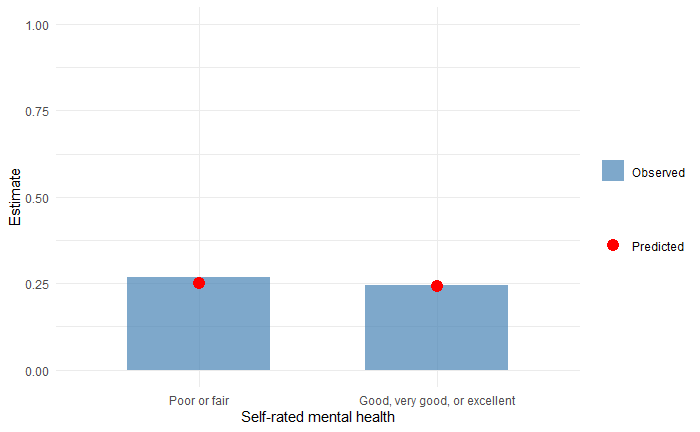

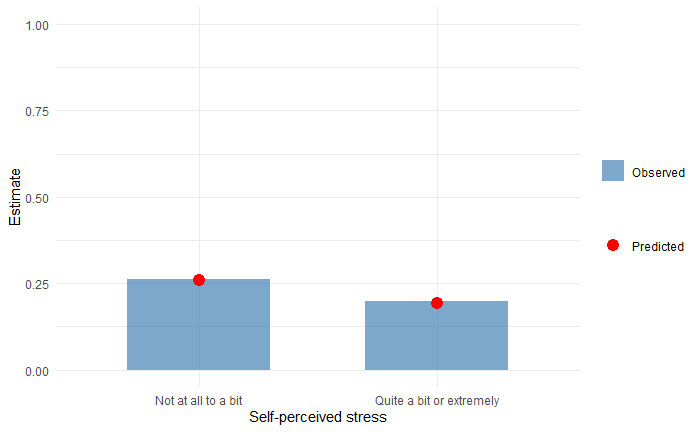

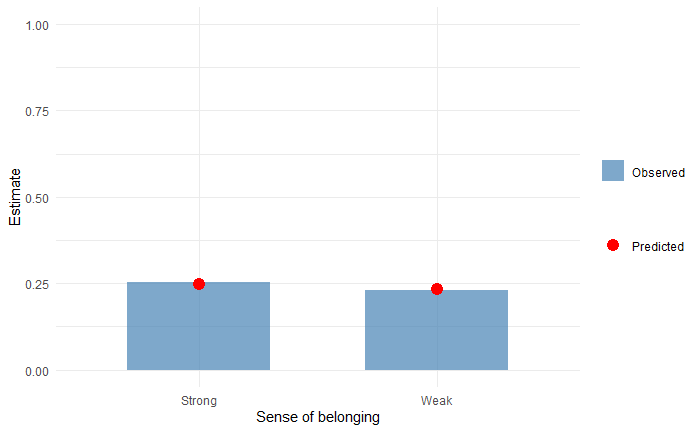

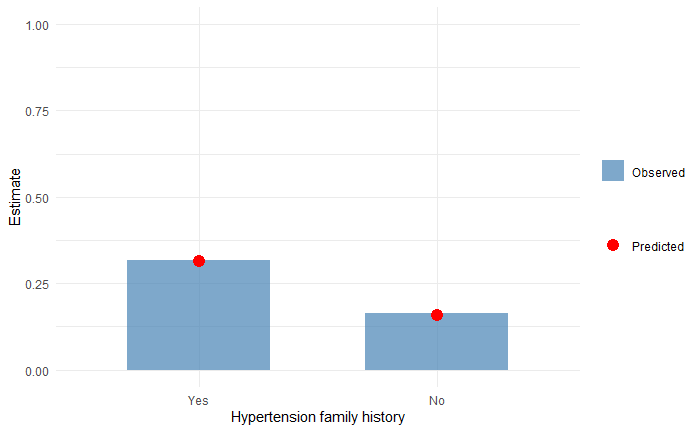

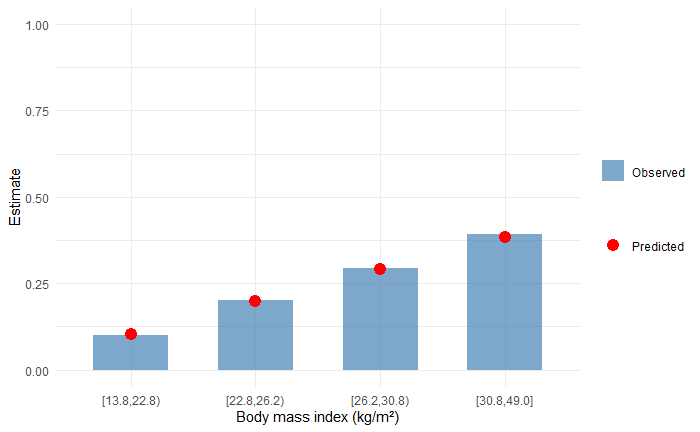

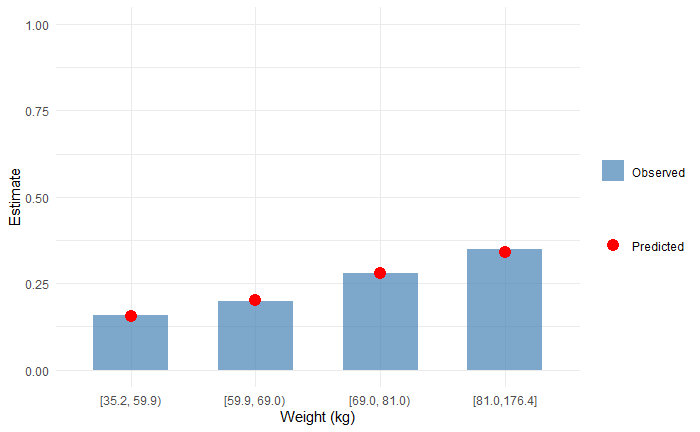

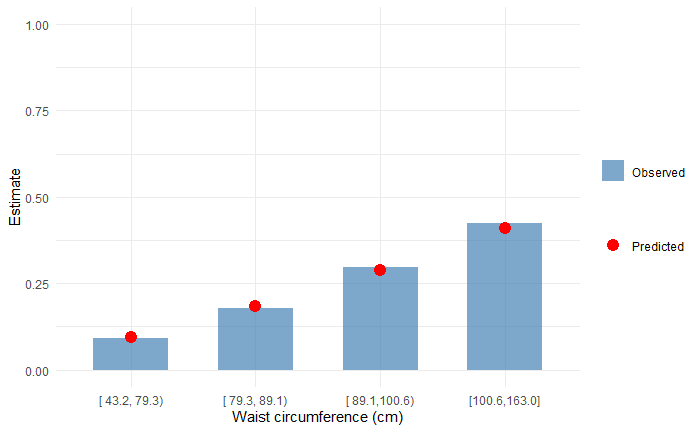

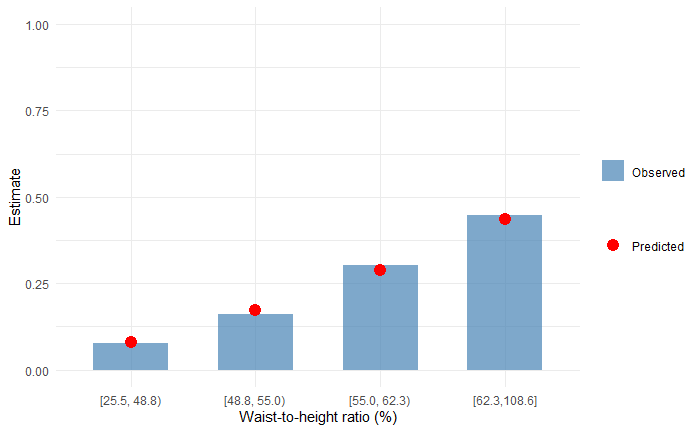

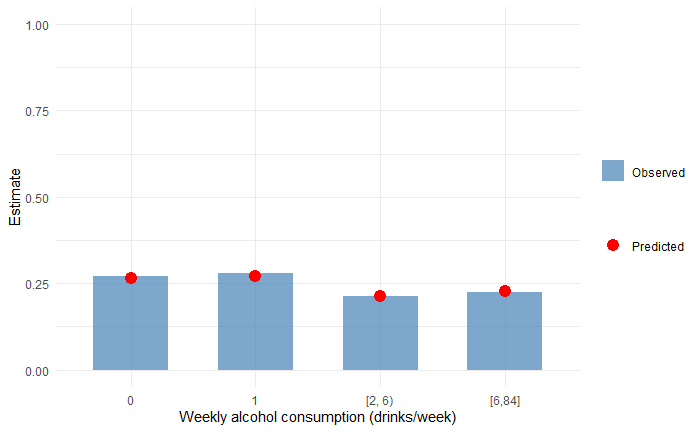

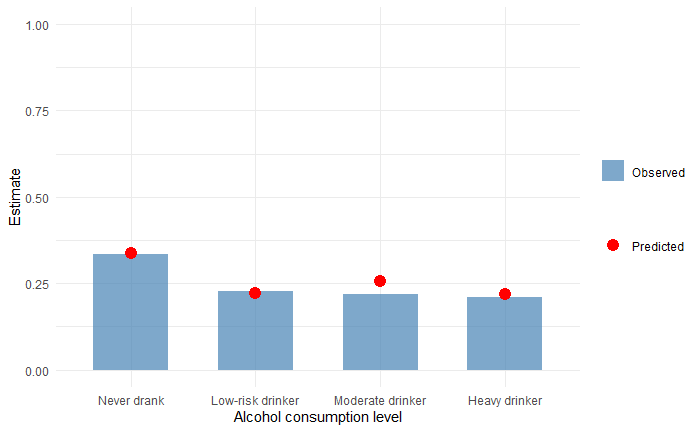

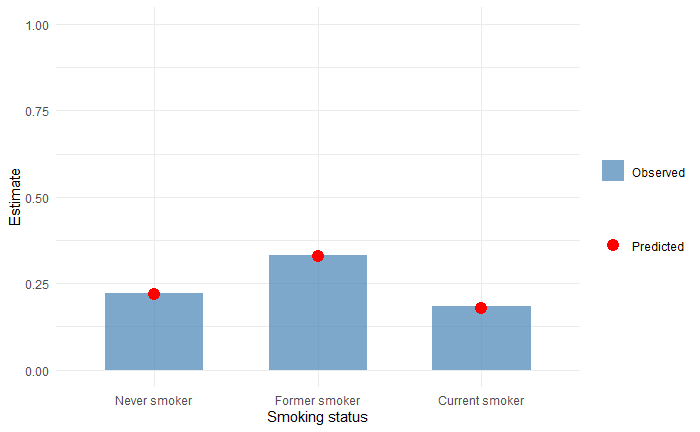

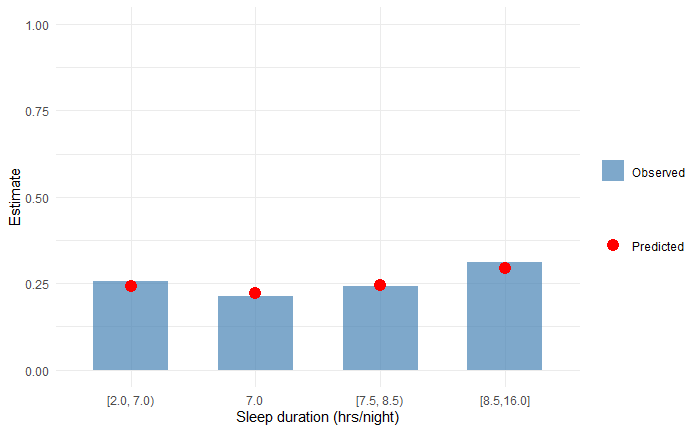

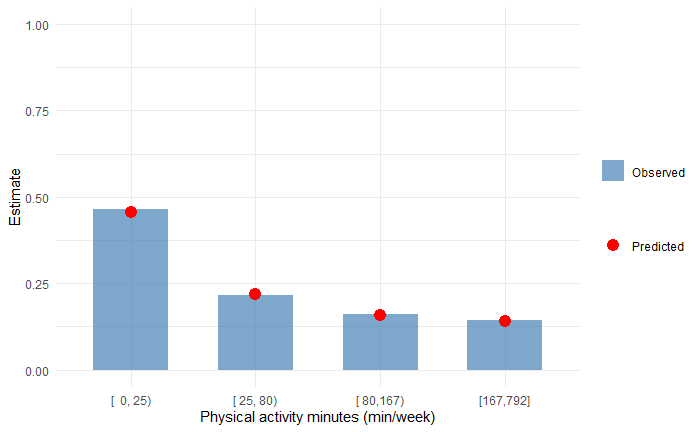

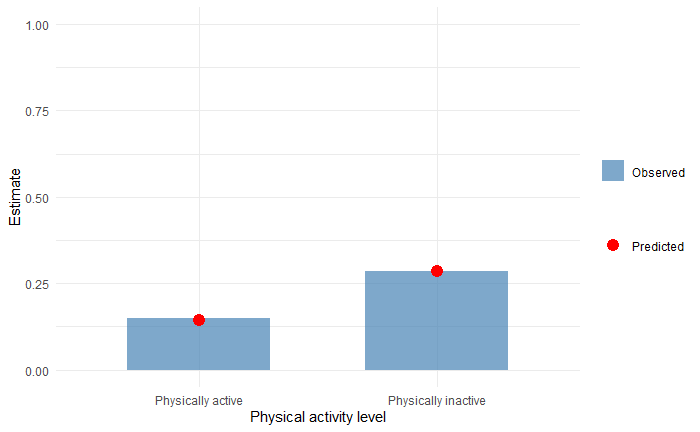

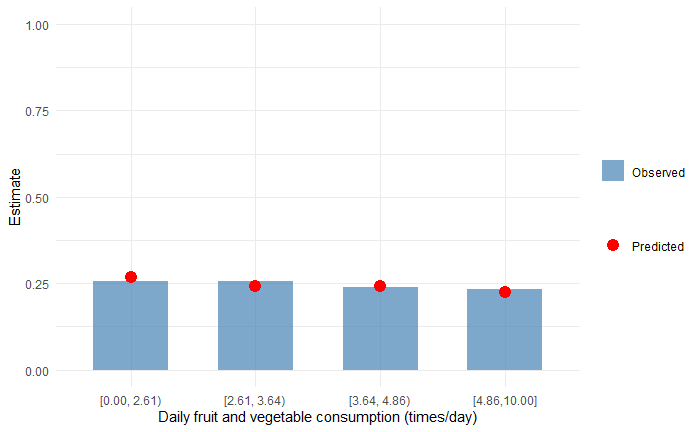

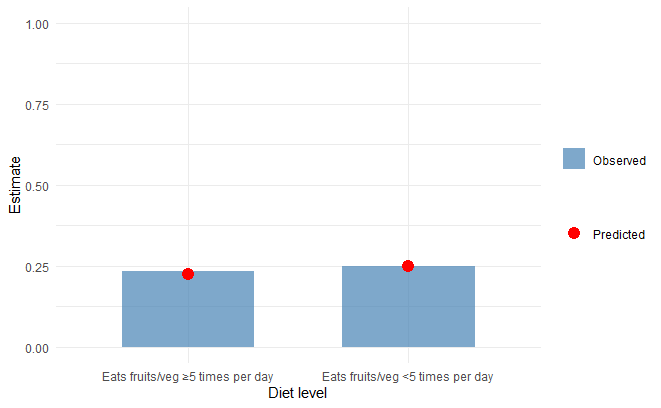

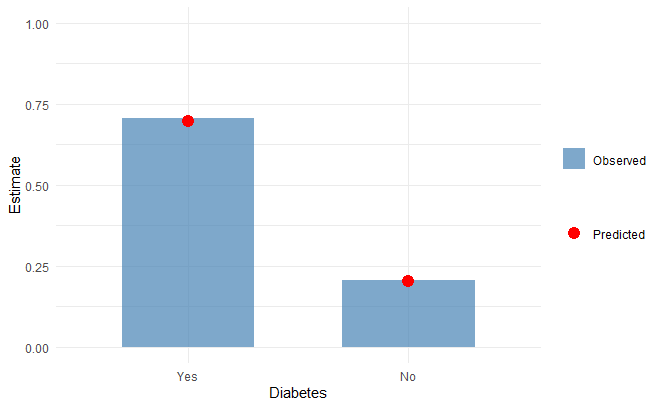

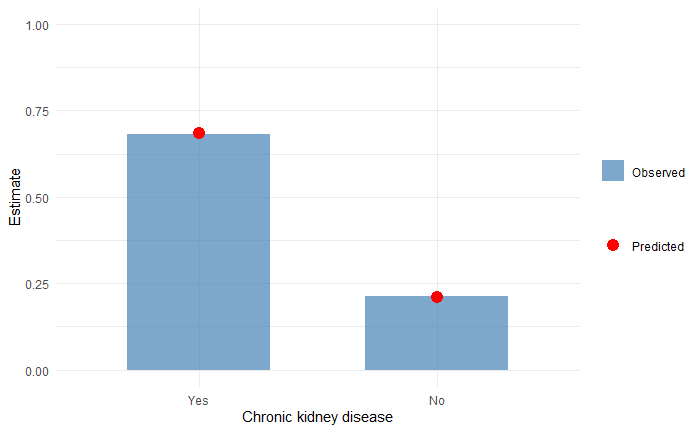

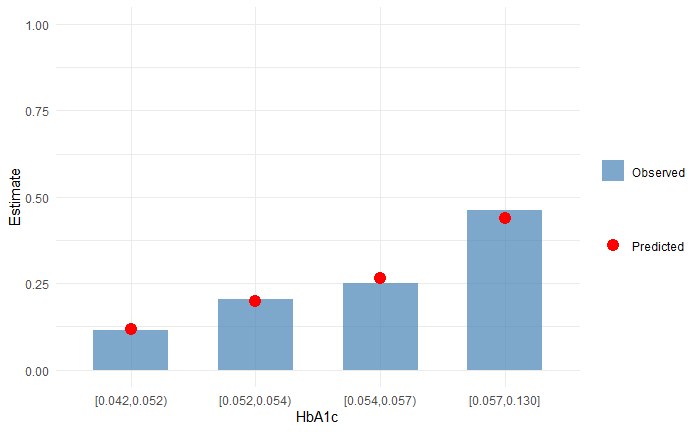

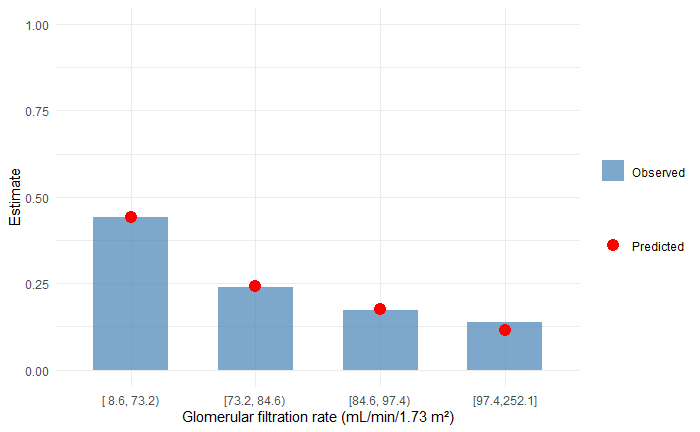

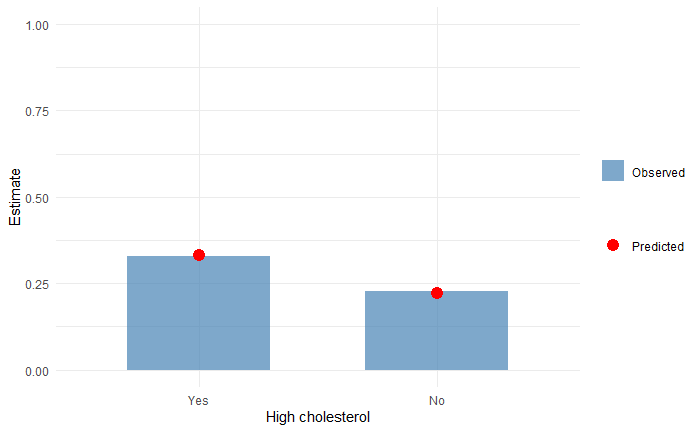

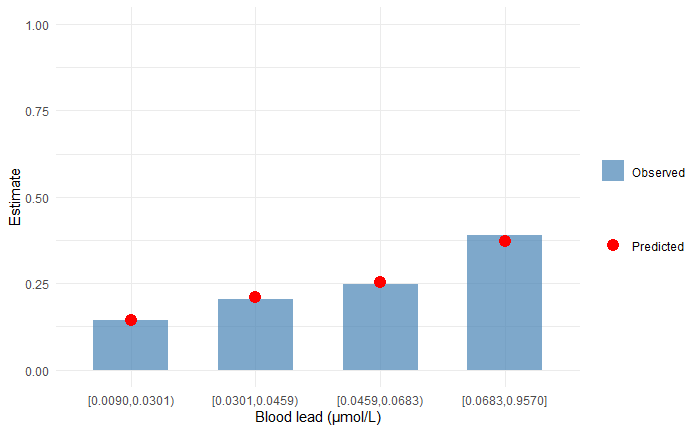

### **Appendix 7 – Calibration across policy-relevant subgroups for reduced models**

**Male reduced model:**

**X – excluded subgroup with observed estimate less than 5%**

*** – subgroup with difference between observed and predicted estimates over 20%**

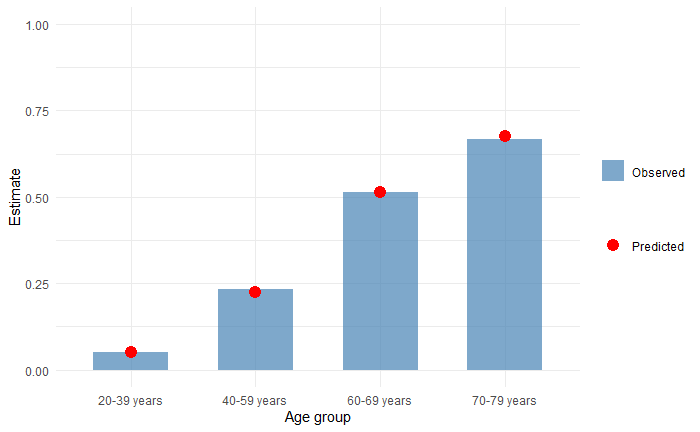

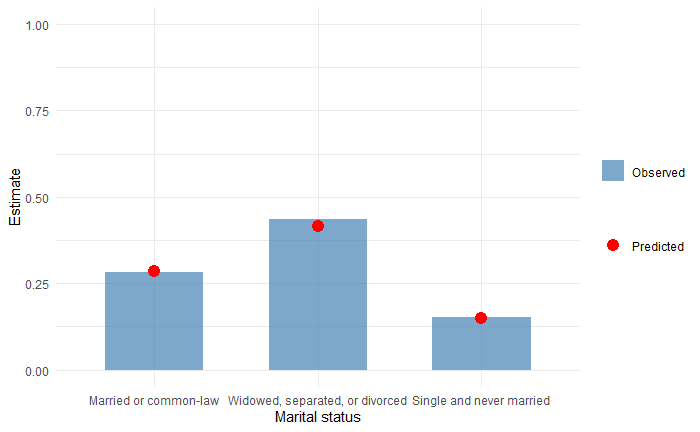

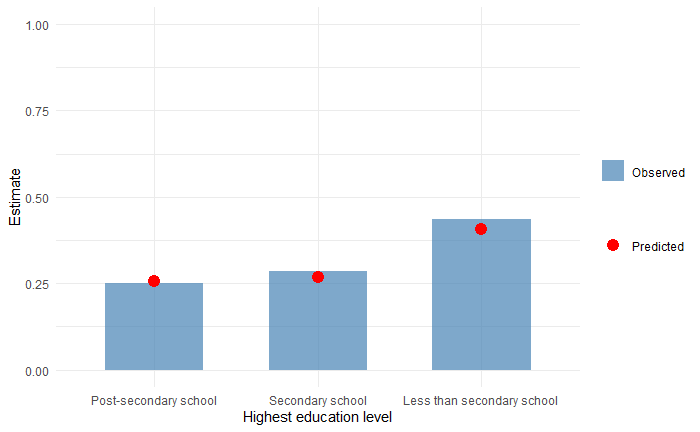

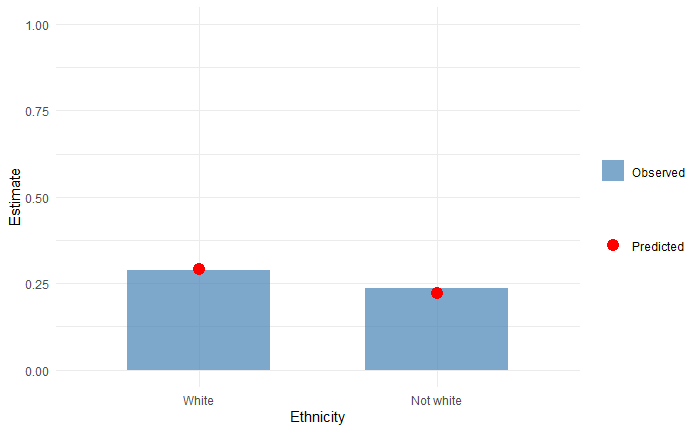

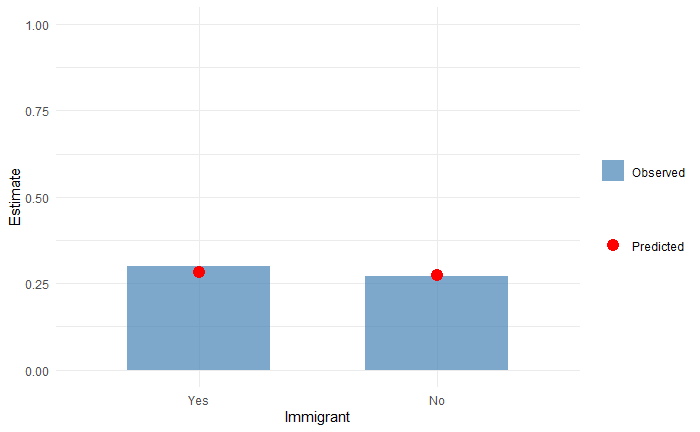

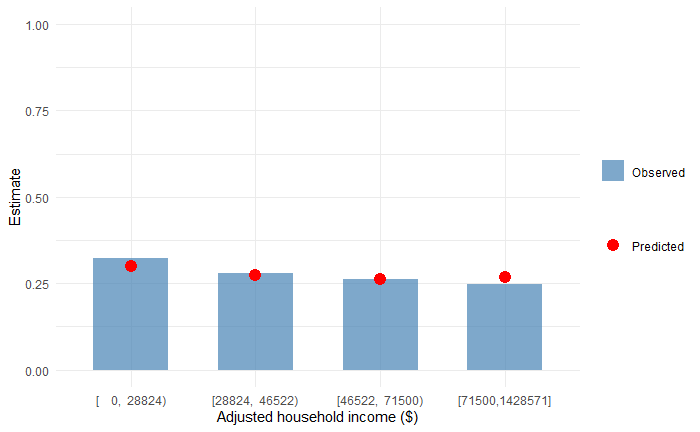

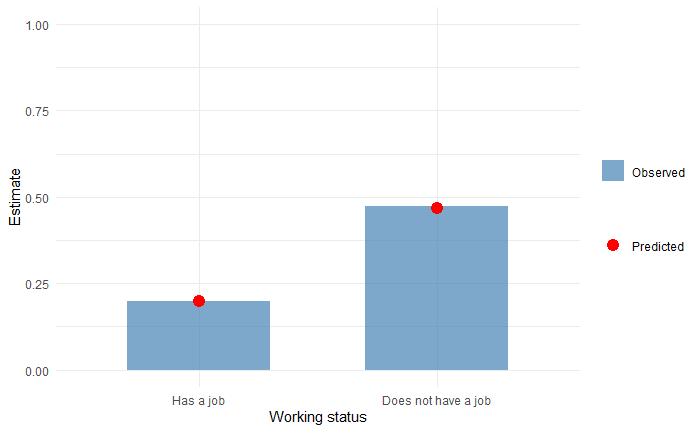

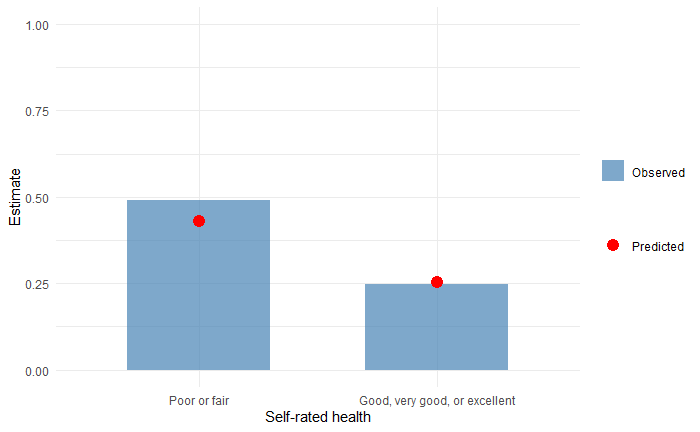

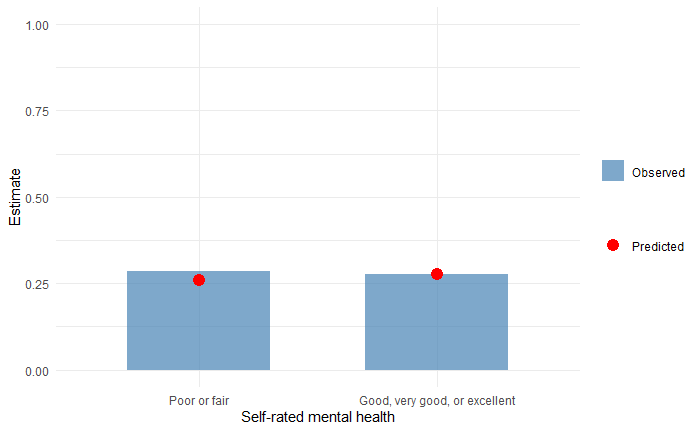

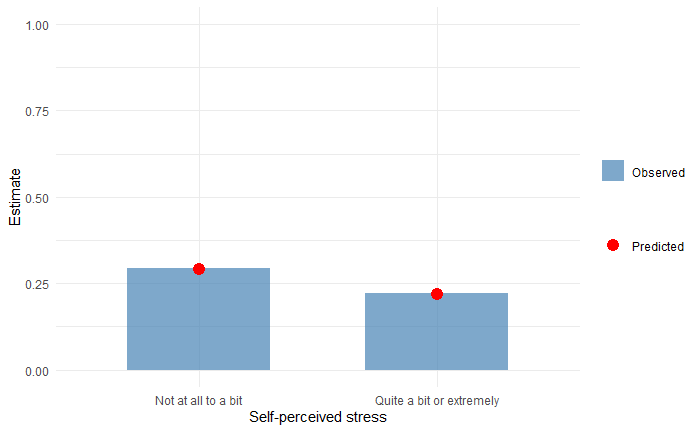

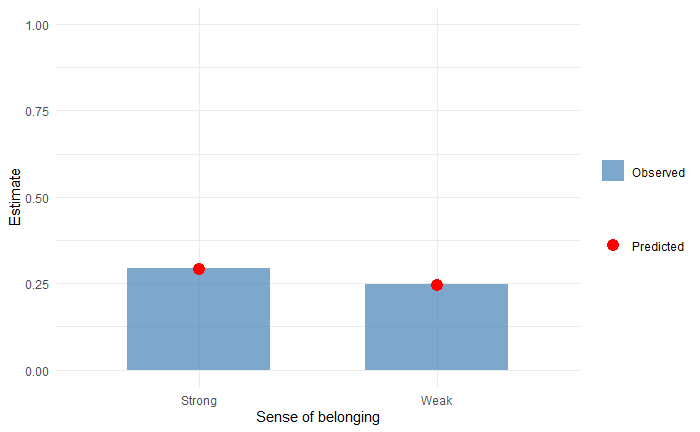

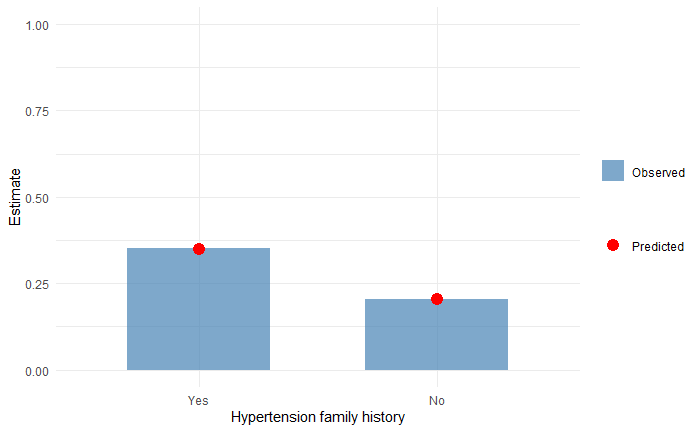

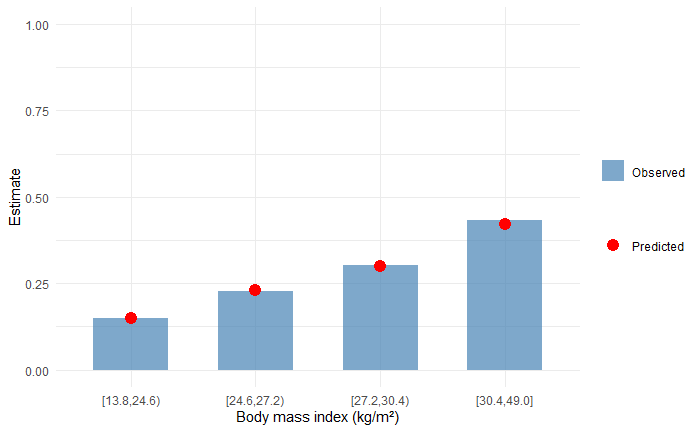

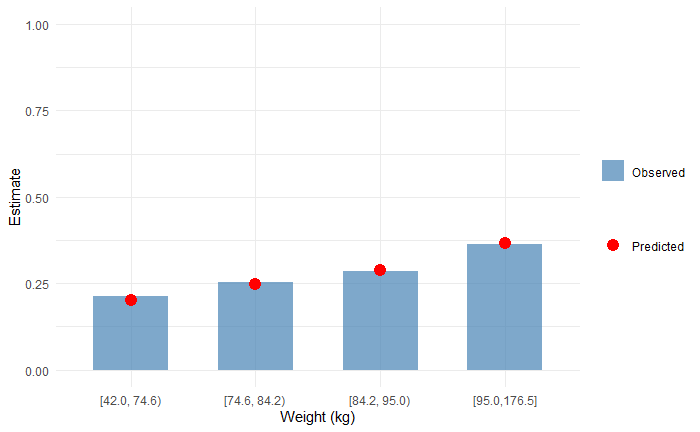

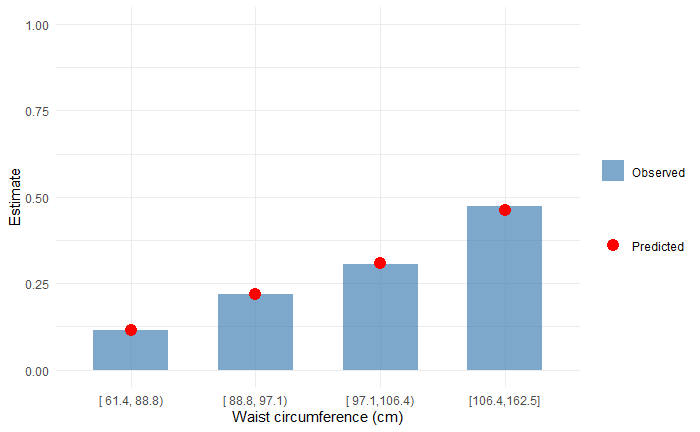

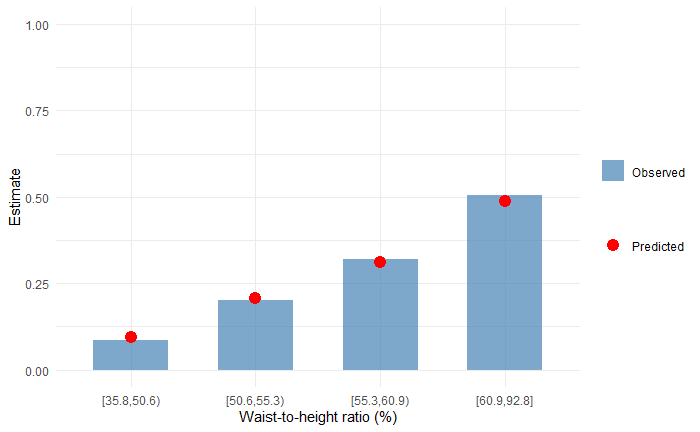

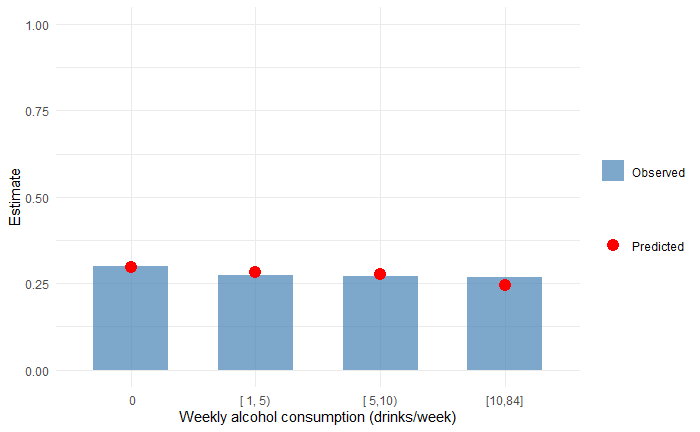

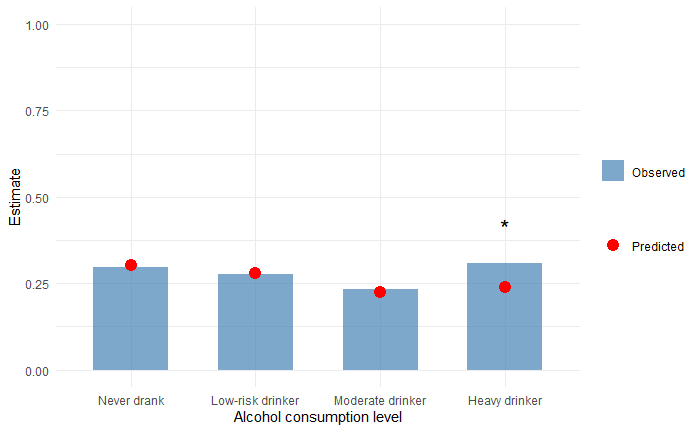

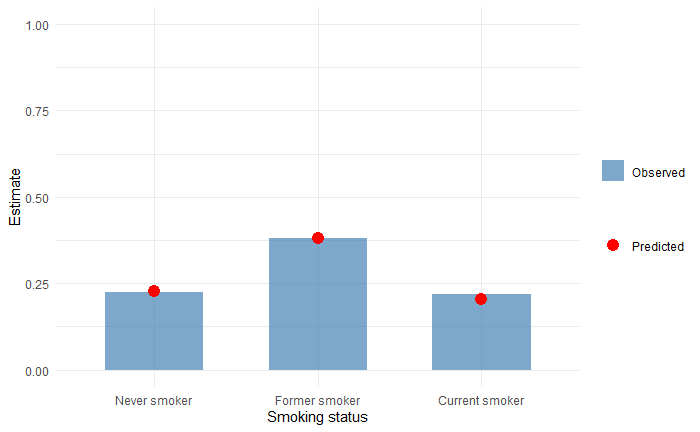

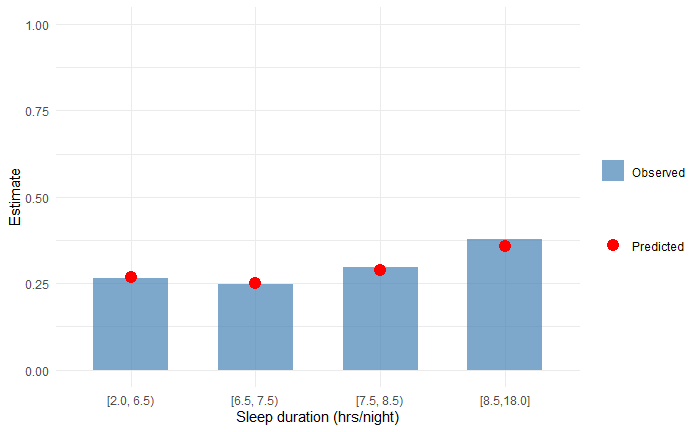

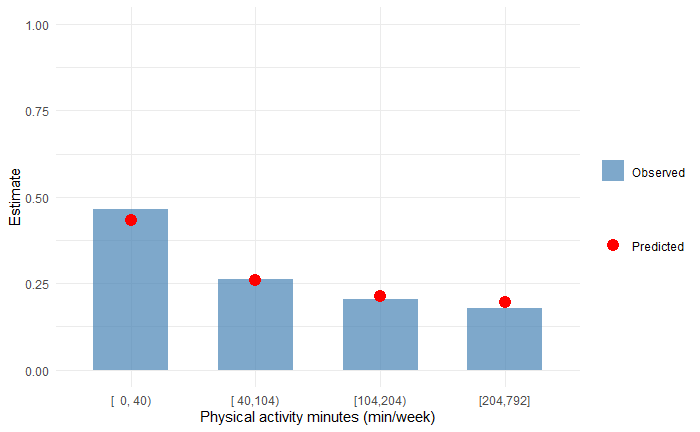

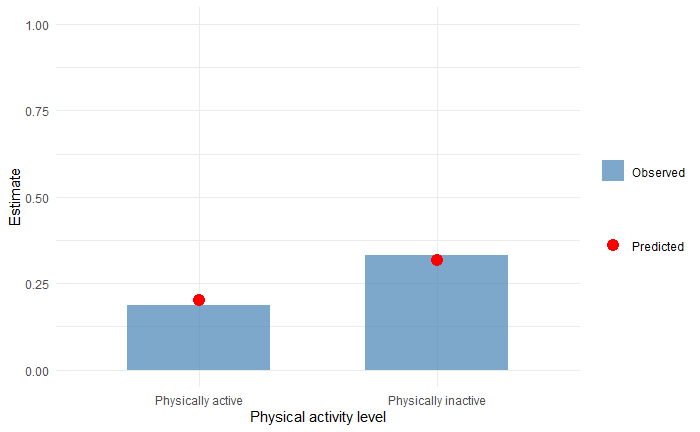

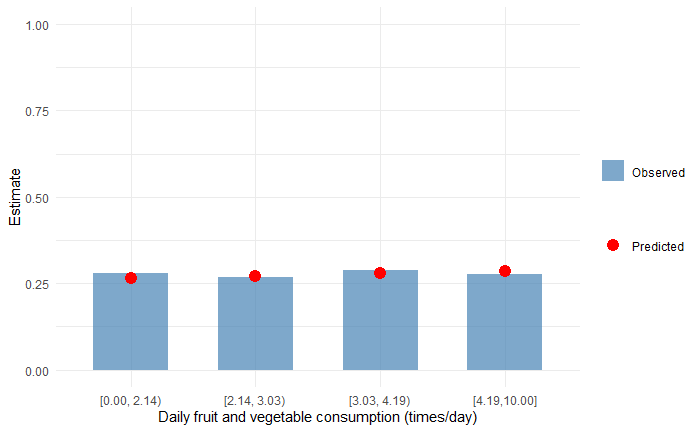

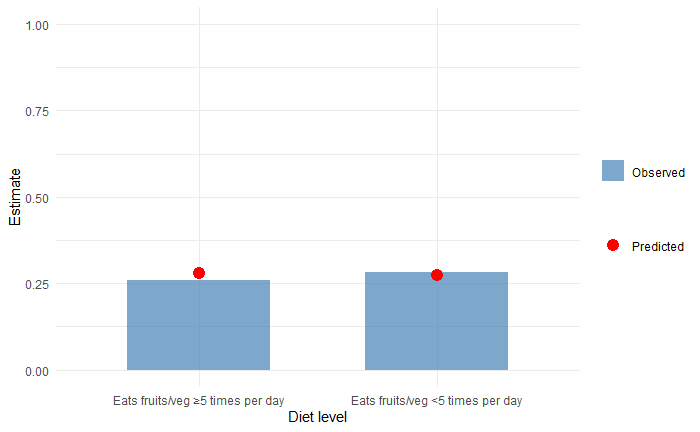

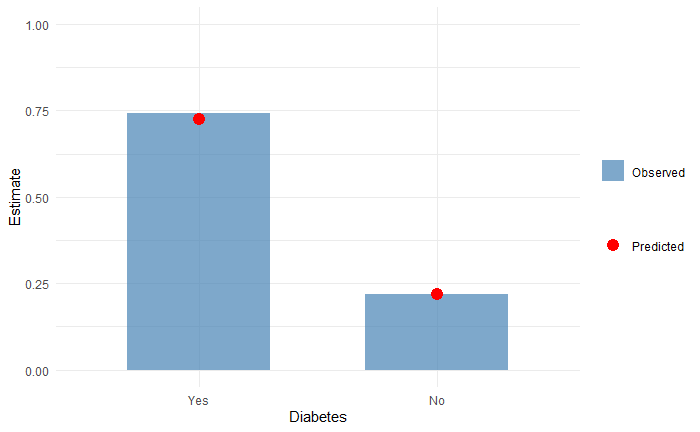

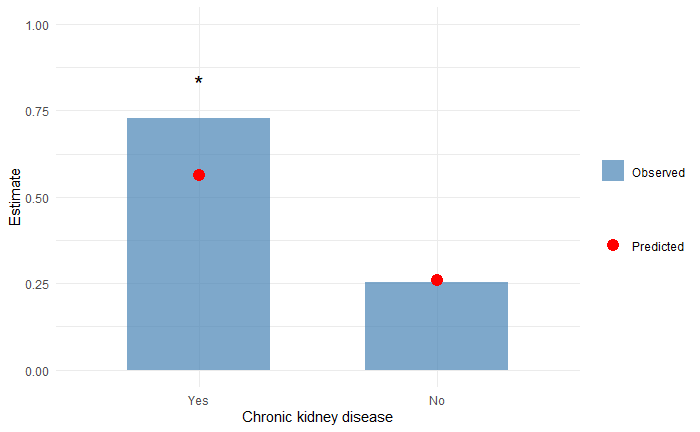

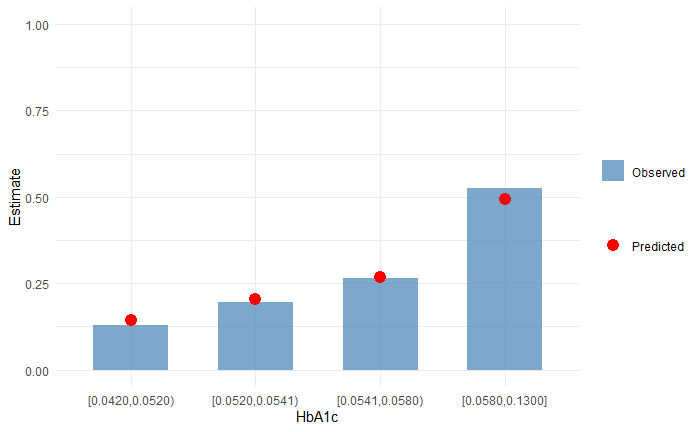

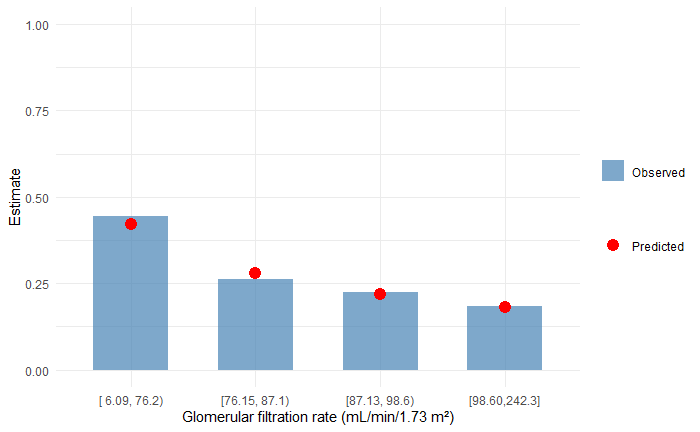

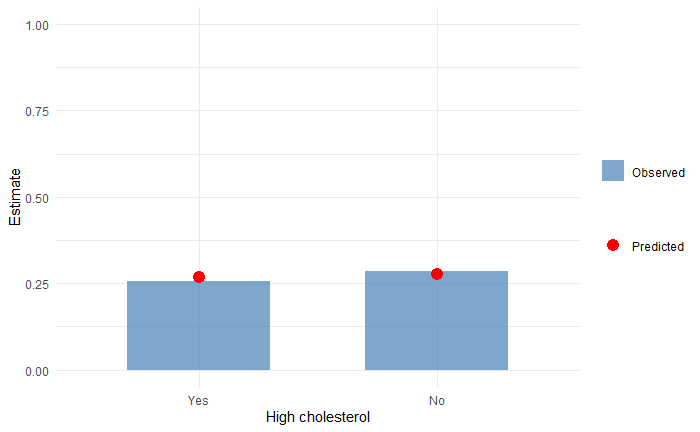

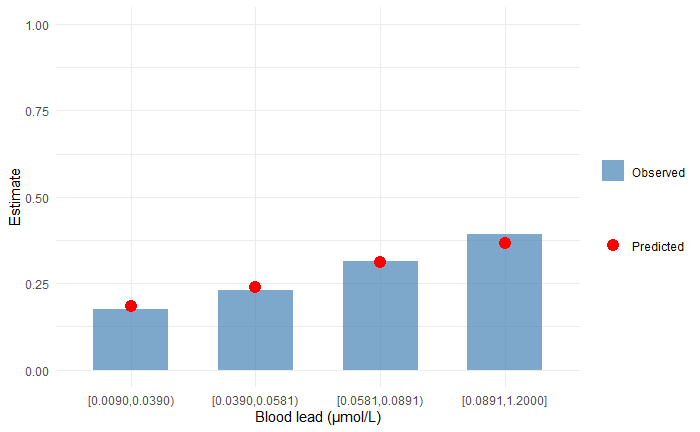

**Female reduced model:**

**X – excluded subgroup with observed estimate less than 5%**

*** – subgroup with difference between observed and predicted estimates over 20%**

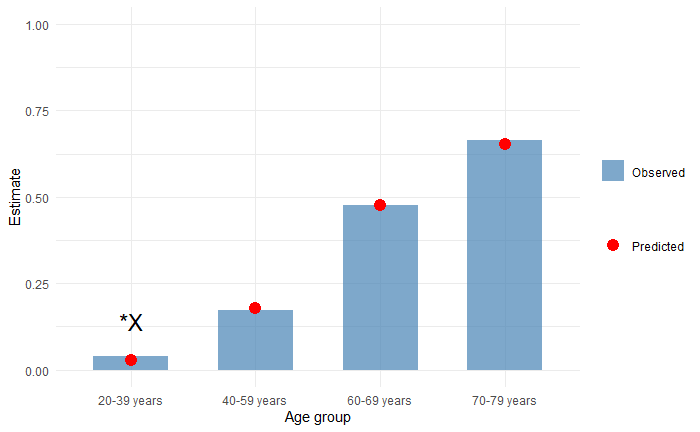

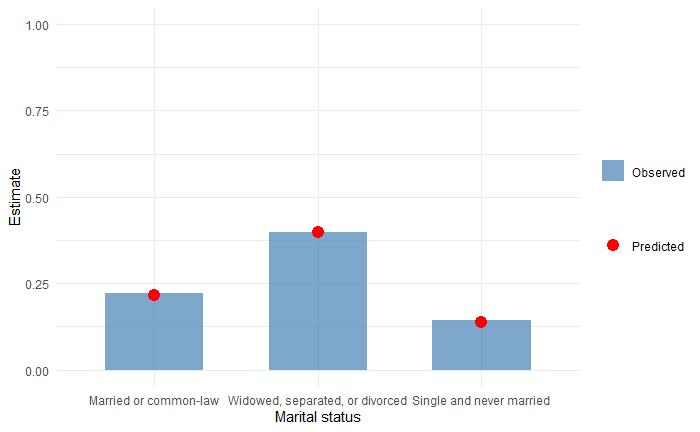

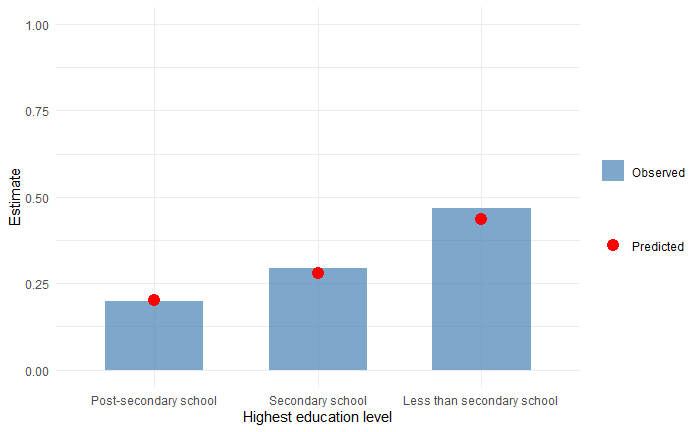

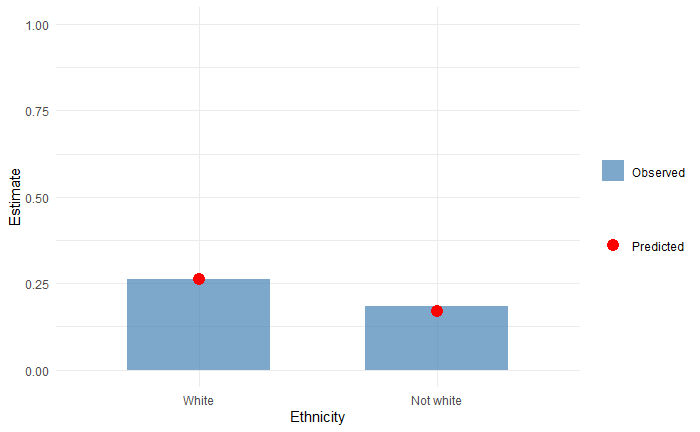

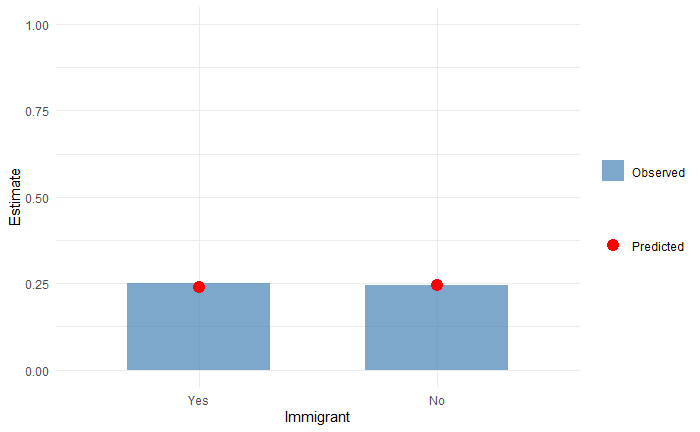

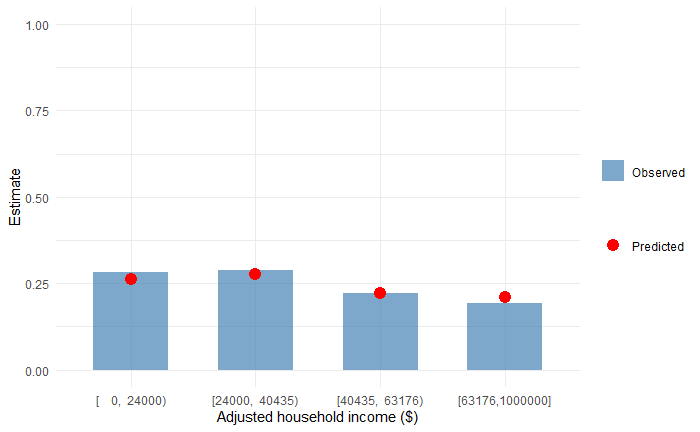

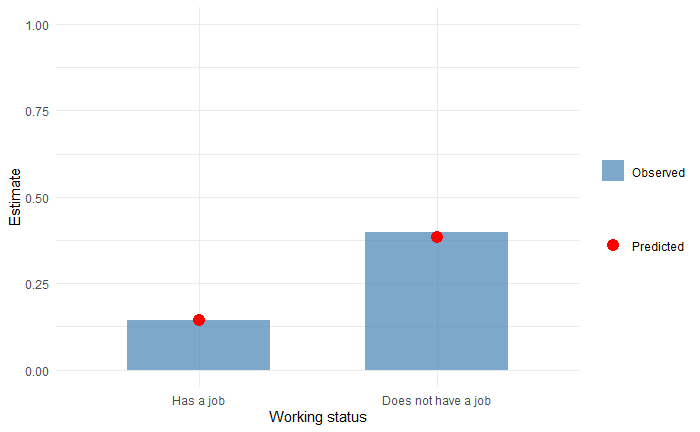

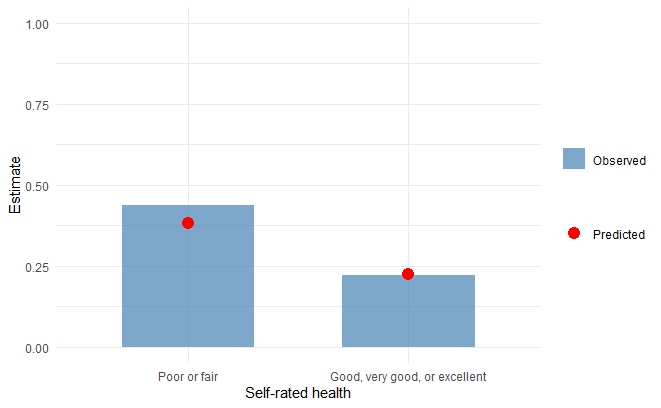

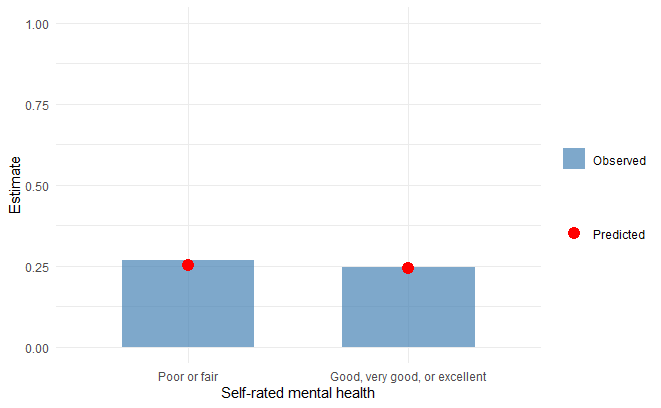

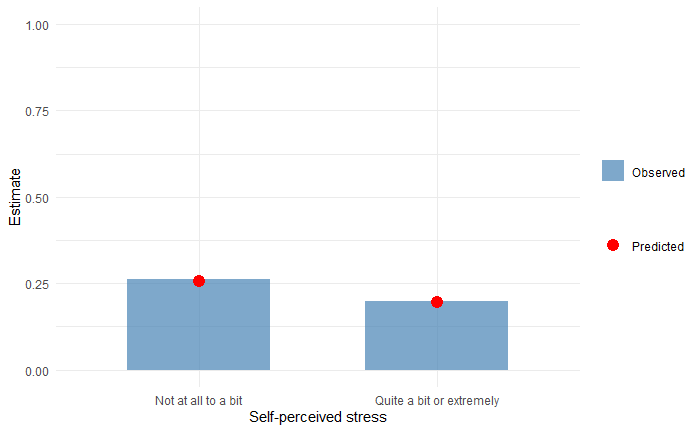

### **Appendix 8 – Sensitivity analyses**

**Calibration plots of models dropping respondents with missing data:**

Male full model:

Female full model:

Male reduced model:

Female reduced model:

**Calibration plots of models derived from four imputed datasets by same MICE model:**

Male full model:

Female full model:

Male reduced model:

Female reduced model:

**Calibration plots of models leaving skewness:**

Male full model:

Female full model:

Male reduced model:

Female reduced model:

**Calibration plots of models with waist-to-height ratio and its interactions re-included:**

Male full model:

Female full model:

Male reduced model:

**Calibration plots of models with linear interactions:**

Male full model:

Female full model:

Male reduced model:

Female reduced model:

**Calibration plots of models for hypertension ascertained with unadjusted blood pressures:**

Male full model:

Female full model:

Male reduced model:

Female reduced model:

**Cumulative predicted probability curves of models for hypertension ascertained with unadjusted blood pressures:**

Male full model:

Female full model:

Male reduced model:

Female reduced model:

**Calibration plots of models excluding hypertensives classified on medications alone:**

Male full model:

Female full model:

Male reduced model:

Female reduced model:

**Cumulative predicted probability curves of models excluding hypertensives classified on medications alone (i.e., those with controlled hypertension):**

Male full model:

Female full model:

Male reduced model:

Female reduced model:

**Calibration plots of models using 130/80 mm Hg blood pressure cutoffs to define hypertension for all respondents:**

Male full model:

Female full model:

Male reduced model:

Female reduced model:

**Cumulative predicted probability curves of models using 130/80 mm Hg blood pressure cutoffs to define hypertension for all respondents:**

Male full model:

Female full model:

Male reduced model:

Female reduced model:
